# Genetic and Epigenetic Dysregulation of CR1 is Associated with Catastrophic Antiphospholipid Syndrome (CAPS)

**DOI:** 10.1101/2025.05.01.25326429

**Authors:** Nikhil Ranjan, Michael Cole, Gloria F. Gerber, Mark A. Crowther, Evan M. Braunstein, Daniel Flores-Guerrero, Kathy Haddaway, Alexis Reed, Michael B. Streiff, MD Keith R. McCrae, Michelle Petri, Shruti Chaturvedi, Robert A. Brodsky

## Abstract

**Objective:** Catastrophic antiphospholipid syndrome (CAPS), characterized by widespread thrombosis and multi-organ failure, is associated with high morbidity and mortality. We previously established complement activation as a pathogenic driver of CAPS and identified rare germline variants in complement-regulatory genes including Complement Receptor 1 (*CR1*) in 50% of CAPS.

**Methods:** We quantified CR1 expression by flow cytometry across hematopoietic cell types. CRISPR/Cas9 genome editing of TF-1 (erythroleukemia) cells was performed to generate *CR1* “knock-out” and “knock-in” lines with patient-specific *CR1* variants. Multiomics analysis was performed to investigate the role of methylation in CR1 expression in patients with reduced CR1 expression. Functional impact of low CR1 expression was assessed by complement-mediated cell killing using modified Ham (mHam) assay, cell-bound complement degradation products through flow cytometry and circulatory immune complexes (CIC) in serum samples through ELISA.

**Results:** CR1 expression in erythrocytes was markedly reduced on CAPS erythrocytes (n=9, 21.80%) compared to healthy controls (HC; n=32, 82.40%), with promoter hypermethylation emerging as a plausible epigenetic mechanism for CR1 downregulation. A novel germline variant (*CR1-*V2125L; *rs202148801*) mitigated CR1 expression and increased complement-mediated cell death of knock-in cell lines. Erythrocytes from the patient with the *CR1-*V2125L variant had low CR1 expression. Levels of CIC, which are bound and cleared by CR1 on erythrocytes, were higher in acute CAPS (n=3, 25.55 µg Eq/ml) than healthy controls (n=3, 7.445 µg Eq/ml). Five patients were treated with C5 inhibition which mitigated thrombosis.

**Conclusion:** Genetic or epigenetic-mediated CR1 deficiency is a potential hallmark of CAPS and predicts response to C5 inhibition.

## 1. Introduction

Antiphospholipid syndrome (APS) is a thromboinflammatory disorder characterized by arterial or venous thrombosis and/or recurrent fetal loss in the presence of persistent antiphospholipid antibodies (aPL), namely lupus anticoagulant (LA), anticardiolipin (aCL) and/or anti-β2 glycoprotein-I antibodies (anti-β2GPI)[1, 2]. The rate of recurrent thrombosis in APS may exceed 20% within the first two years despite anticoagulation[3–5]. “Catastrophic” APS (CAPS) is a rare subset (∼1-2%) of APS, characterized by rapidly developing widespread thrombosis and multi-organ failure and is associated with high morbidity and mortality[1, 6]. Currently, no biomarkers predict the risk of developing CAPS, and it is unclear what differentiates APS and CAPS from a biological perspective. Indefinite anticoagulation with a vitamin K antagonist is the standard of care for thrombotic APS; however, this treatment is suboptimal for some patients with anticoagulant-refractory thrombosis and those with CAPS.

Complement is implicated in the pathophysiology of thrombosis in CAPS and complement inhibition has been successfully used to treat anticoagulant refractory thrombosis in APS and CAPS[7–10]. We demonstrated abnormal complement activation in >85% of CAPS sera[11, 12] and found that 48% (9/19) of CAPS patients had rare germline variants in complement regulatory genes, a rate that is comparable to atypical hemolytic uremic syndrome, a prototypical complement-mediated thrombotic disorder[13, 14]. Of these, 44% (4/9) harbored novel rare Single Nucleotide Variants (SNVs) variants in Complement Receptor 1 (*CR1*).

*CR1* encodes a ∼160-300kDa transmembrane glycoprotein expressed on neutrophils, B lymphocytes, erythrocytes and monocytes but not on platelets [15–18]. CR1 regulates complement through C4b and C3b binding and plays a dominant role in the clearance of soluble immune complexes[19],[20–22]. Similar to complement factor H (CFH), CR1 possesses decay-accelerating activity, leading to the dissociation of the C3 or C5 convertase, and acts as a cofactor for complement factor I (CFI) in the cleavage of C4b and C3b[22, 23],[19, 20, 24, 25]. Erythrocytes express 100-1000 CR1 molecules per cell [26]. CR1 also carries the Knops blood group system, which currently includes 13 antigens with 5 antithetical antigen pairs (Kn^a^/Kn^b^, McC^a^/McC^b^, Sl^a^/Vil, KCAM/KDAS, DACY/YCAD) [27]. The Helgeson phenotype, previously considered to be the Knops "null" phenotype, is serologically defined as Kn(a-) McC(a-) Sl(a-) Yk(a-) and is characterized by extremely low CR1 expression on erythrocytes (<100 CR1 molecules per cell) [27, 28]. SNVs in the *CR1* gene are linked to reduced CR1 expression. Previously described variants associated with the Helgeson phenotype include rs11118133: A > T, rs2274567: A > G (represents the DACY-YCAD+ phenotype) and rs3811381: C > G[26, 29–31]. A recent study identified a SNV on *CR1* (rs11117991: T > C) that disrupts a GATA1-binding site and reduces CR1 expression on erythrocytes[32]. GATA1 is a transcription factor crucial for regulating gene expression in erythroid cells, however *CR1* expression in granulocytes (neutrophils, basophils and eosinophils) and monocytes are governed PU.1 [33] [16, 34].

Here, we demonstrate patients presenting clinically with CAPS have reduced CR1 expression, consistent with Helgeson-like phenotype. This reduced expression may be attributed to germline mutations or hypermethylation in *CR1*. CR1 deficiency leads to impaired clearance of immune complexes, complement dysregulation, and a severe thrombotic phenotype mitigated by C5 inhibition *in vitro* and *in vivo*. We propose that CR1 deficiency will help identify patients at the highest risk of refractory APS or CAPS and those likely to benefit from complement inhibition.

## Materials and methods

### Patient classification and samples

We included patients with APS or CAPS enrolled in the Johns Hopkins Complement Associated Disorders Registry (CADR) as well as healthy controls without known APS or thrombosis. Patients were enrolled in the registry based on the International Society on Thrombosis and Hemostasis criteria for APS[2]. All CAPS patients and the majority of patients in the APS cohort without a history of CAPS also met American College of Rheumatology (ACR)/ European Alliance of Associations for Rheumatology (EULAR) 2023 criteria for APS[35]. Recurrent thrombosis was confirmed by imaging at any time before enrollment or during follow-up in the registry. For patients with multiple tests for aPLs, we report the results drawn closest to the study sample. Definite or probable CAPS was diagnosed according to international consensus criteria, including involvement of 3 or more organs, development of manifestations within a week, histologic confirmation of small vessel thrombosis, and laboratory confirmation of the presence of aPLs [36]. Definite CAPS requires all four criteria while probable CAPS is diagnosed if 3 criteria are met [36]. We included patients with both definite and probable CAPS because the outcomes of patients with probable CAPS are comparable to those of patients with definite CAPS[37] (Table 1). Clinically, it is oftentimes challenging to obtain a tissue biopsy to confirm microvascular thrombosis, resulting in many patients having “probable” CAPS.

**Table 1.** Clinical features for CAPS/ probable CAPS patients.

| Patient | Age, sex | APS Laboratory Criteria<sup>a</sup> | Thrombotic and obstetric history | Thrombotic Trigger | CAPS Manifestations | CAPS treatment | Current Treatment | CR1 % expression |
| --- | --- | --- | --- | --- | --- | --- | --- | --- |
| P1 | 36-40, F | Triple positive; primary | HELLP x3, fetal death x2, PVT, renal failure VTE on anticoagulation | Pregnancy | Renal failure, thrombocytopenia, and elevated transaminases | plasma exchange, high-dose corticosteroids, anticoagulation | Anticoagulation, no further TE outside pregnancy | 15.2 |
| P2 | 41-45, M | Triple positive; primary | Stroke, biopsy-proven thrombotic vasculopathy of toes | None | Transient ischemic attack, myocardial infarction and diffuse alveolar hemorrhage | Anticoagulation, antiplatelet agents, corticosteroids, rituximab, C5i | Continues on C5i, anticoagulation stopped, no further TE | 8.4 |
| P3 | 51-55, M | Triple positive; secondary | DVT, PE, myocardial infarction | Infection | Thrombocytopenia, liver injury, proteinuria, and encephalopathy, bilateral adrenal hemorrhage | Anticoagulation, hydroxychloroquine, plasma exchange, corticosteroids, rituximab, C5i | Continues on C5i and anticoagulation, no further TE | 1.9 |
| P4 | 56-60, F | LAC; secondary | Recurrent PE on anticoagulation, multiple episodes of ischemic colitis, adrenal infarction, ischemic retinopathy and TIA | None | Pulmonary embolism, TIA, ischemic colitis, ischemic retinopathy | Anticoagulation, antiplatelet agents, hydroxychloroquine, rituximab, C5i | Continues on C5i; anticoagulation stopped, 1 DVT post-op in the setting of delayed C5i | 23 |
| P5 | 21-25, F | LAC; secondary | Recurrent DVT (both on and off anticoagulation), placental abruption, chronic thrombocytopenia, HELLP syndrome | Pregnancy |  |  | Anticoagulation, hydroxychloroquine | 20.9 |
| P6 | 56-60, M | Triple positive; primary | Recurrent DVT on anticoagulation, IVC filter thrombosis, renal vein thrombosis, stroke | None |  |  | Anticoagulation, aspirin, hydroxychloroquine | 27.2 |
| P7 | 56-60, M | LAC, aCL IgM; primary | SMA thrombosis, SMV thrombosis, PVT | Surgery, infection | Progressive SMV thrombosis, bilateral adrenal hemorrhage, stroke, splenic infarcts | IVIg, plasma exchange, anticoagulation, C5i | Anticoagulation, C5i | 15.9 |
| P8 | 51-55, M | Triple positive, primary | LLE arterial thrombosis | None | Thrombocytopenia. DAH, acute renal failure with renal TMA | Plasma exchange, steroids, anticoagulation and IVIg, C5i | Continues on C5i, hydroxychloroquine, warfarin and clopidogrel | 56.3 |
| P9 | 21-25, M | Triple positive, primary | Recurrent venous thrombosis including cerebral vein thrombosis, bilateral leg arterial thrombosis | Non-compliance with anticoagulant | Bilateral limb-threatening leg ischemia | Thrombotic storm treated with prednisone, plasmapheresis and unfractionated heparin transitioned to fondaparinux | Anticoagulation, aspirin, hydroxychloroquine | 27.4 |

^Primary APS is defined in the absence of a second underlying autoimmune disorder, commonly lupus, whereas secondary APS occurs in the setting of a concomitant diagnosis of a defined autoimmune disorder.
HELLP, Hemolysis, Elevated Liver enzymes, and Low Platelets; PVT, Portal Vein Thrombosis; VTE, Venous Thromboembolism; C5i, C5 inhibitor; TE, Thromboembolism; LAC, Lupus anticoagulant; aCL, Anti-cardiolipin antibodies; a β2GPI, Anti β2 Glycoprotein-I antibodies; PE, Pulmonary Embolism; DVT, Deep Vein Thrombosis; TIA, Transient Ischemic Attack; IVC, Inferior Vena Cava; SMA, Superior Mesenteric Artery; SMV, Superior Mesenteric Vein; LLE, left lower extremity; DAH, Diffuse Alveolar Hemorrhage; TMA, Thrombotic Microangiopathy; IVIg, Intravenous Immunoglobulin

Blood was collected by venipuncture in EDTA and serum separation tubes. Serum was allowed to clot prior to centrifugation at 4°C then separated and stored at −80°C. Whole blood was used to isolate genomic DNA for sequencing. This study was approved by the Institutional Review Board at Johns Hopkins University and was conducted in accordance with the Declaration of Helsinki. All patients provided written informed consent and were not involved in the design, conduct, reporting, or dissemination plans of this research.

### CR1 expression by flow cytometry

Whole blood or Ammonium-Chloride-Potassium (ACK) lysed cells were washed, suspended in Hanks Balanced Salt Solution (HBSS)+1%BSA (HBSSA), and stained with fluorescent dye-conjugated CR1 antibody for 30 minutes at room temperature. Flow cytometry was performed using Cytoflex S (Beckman Coulter, CA, USA) and analyzed using FlowJo.

### Targeted sequencing, multi-omics and CRISPR Cas9-mediated gene editing

Genomic DNA was isolated using a DNeasy Blood & Tissue Kit (Qiagen, Germany) and quantified with Qubit fluorometric assay. 50ng of DNA was used for library preparation using a custom-seq panel of 24 complement regulatory genes described in Table S1 (Illumina, USA). To understand the methylation fraction on *CR1*, we performed whole genome sequencing and multiomics using duet multiomics evoC kit (Biomodal, Cambridge, UK).

The CRISPR/Cas9 system was used to generate CR1 knock-out (KO) and knock-in (KI) cell lines through homology-directed repair (HDR). The experiments described were performed on five cell lines – TF-1*^WT^*, TF-1*^CR1-/-^*, TF-1*^V2125L^*, TF-1*^G2109S^*and TF-1*^S1982G^* unless mentioned otherwise.

### Modified Ham (mHam)

mHAM assay was performed with TF-1^WT^, TF-1^CR1-/-^, TF-1^V2125L^, and TF-1*^G2109S^* cells as previously described [12]. NHS was treated as indicated with heat inactivation (HI) for 30 minutes at 56°C to inactivate complement proteins or addition of targeted complement inhibitors. Following incubation of the cells, serum viability was measured using a WST-1 dye.

### Bioluminescent mHam (bio-mHam)

Bio-mHam was performed as previously described [37]. Bioluminescent HEK293*^PIGA-/-^* cells were incubated with patient serum after respective treatment (heat inactivation and complement inhibitors). Luminescence was monitored serially and percent relative luminescence at 1 hour was calculated as the luminescence of cells treated with the patient serum compared to cells treated with the sample’s heat-inactivated control. A relative 1 h luminescence of ∼12% defines the positive threshold for abnormal complement activity, corresponding to the lower 85th percentile.

### Complement fragment deposition through flow cytometry

Each cell line was incubated with NHS at 37°C for various durations between 5 and 90 minutes). Cells were washed with HBSSA and stained with C3c and C3dneo murine mAbs (Quidel, San Diego, CA) at 5 µg/ml for 30min at 4°C followed by anti-mouse AF488 (Invitrogen, MA, USA) antibody for 30 min at 4°C. Cells were washed and resuspended in HBSSA for flow cytometry analysis using a Cytoflex S flow cytometer (Beckman Coulter, CA, USA). Data was collected from at least 10,000 cells and analyzed using FlowJo.

### Quantification and statistical analysis

Descriptive statistics and analysis were performed using Prism10 (GraphPad, MA, USA). A complete description of experimental details can be found in the supplemental data.

## Results

### Patient cohort and samples

A total of 11 patients with CAPS, 8 with thrombotic APS, and 32 healthy controls were evaluated. Clinical presentation and characteristics of the CAPS group are detailed in Table 1. Amongst the 11 patients with CAPS, CR1 sequencing was completed in 8 patients (three had not consented to genetic studies), and erythrocytes to evaluate CR1 expression were available for 9 (the other two were deceased).

### Loss of CR1 makes APS catastrophic

Building on previous findings that revealed novel germline variants in 48% of CAPS patients, we evaluated CR1 expression in our CAPS cohort. CR1 expression was significantly reduced in patients with CAPS (n=9, 21.80% ± 5.146) compared to the patients with APS (n=8, 85.30% ± 2.797) and healthy controls (n=32, 82.40% ± 2.291). The mean erythrocyte CR1 expression for the Helgeson phenotype controls (obtained from the Johns Hopkins transfusion medicine division, n=2) was 24.80% ± 7.1 (Figure 1A-B).

**Figure 1.**
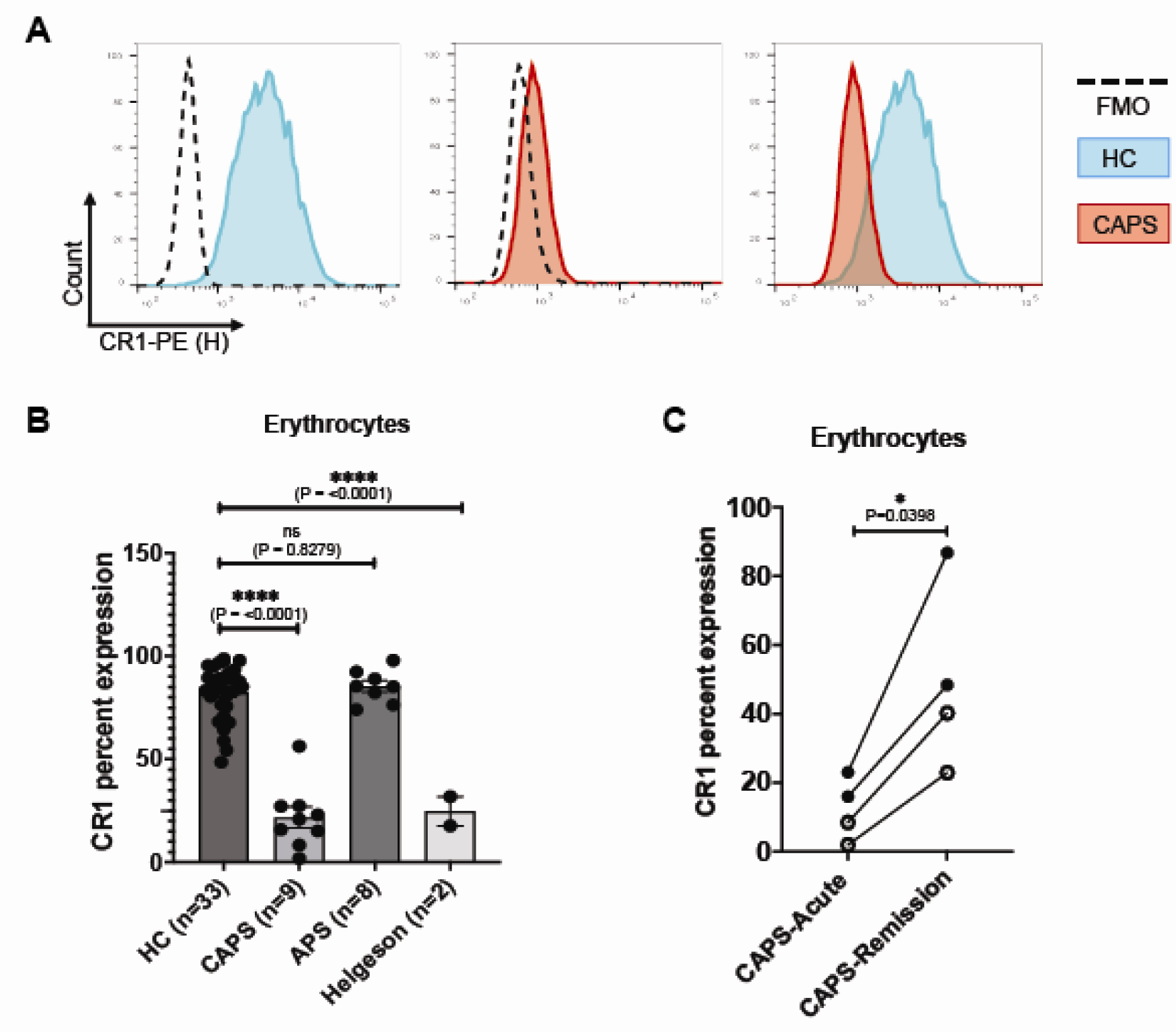
Loss of CR1 impairs immune clearance and makes APS catastrophic. **A.** Representative histograms of CR1 expression in whole blood samples from healthy control (blue) or CAPS patient (red). Dashed line represents fluorescence minus one (FMO) control. **B.** CR1 expression in erythrocytes from HC (82.40% ± 2.291, n=32), CAPS (21.80% ± 5.146, n=9), APS (85.30% ± 2.797, n=8) and Helgeson phenotype (24.80% ± 7.1, n=2). **C.** CR1 expression in serial samples from CAPS patients representing increased CR1 expression during the remission phase (49.58% ± 13.50) of the disease as compared to the acute phase (12.30% ± 4.572). Patients identified with Helgeson phenotype (rs11117991) are represented as hollow circles. Data is represented as mean ± SEM, ns: not significant, ****: P < 0.0001.

Serial samples from CAPS patients P2, P3, P4 and P7 were collected during the acute and remission phase of CAPS. Samples collected during the remission phase showed significantly higher CR1 expression (49.58% ± 13.50) compared to the acute phase (12.30% ± 4.572), suggesting a loss of CR1 during the acute phase of the disease (Figure 1C).

### *CR1* Polymorphisms and patient-specific cell line models

*CR1* variants were found in 4 of 8 CAPS patients who underwent sequencing. None of the sequenced control samples (healthy control, n=7 and APS, n=8) had *CR1* variants. The index patient, first identified with a CR1 variant in our cohort was a female in her mid 30’s (P1) with APS (lupus anticoagulant and high titer anticardiolipin IgG) with multiple venous and arterial thrombotic events despite anticoagulation and three pregnancy losses was evaluated during her fourth pregnancy. The mHam assay was negative at 17 weeks gestation, however, the mHam assay became positive at 23 weeks (44.5% cell killing) (Figure 2B). Two days later, she developed thrombocytopenia, liver function tests elevation and hypoxemic respiratory failure consistent with either CAPS or HELLP. Targeted sequencing for 24 complement regulatory genes (Table S1), identified a heterozygous missense mutation in *CR1* (*CR1*^V2125L^; rs202148801, Figure 2A), which was verified by Sanger sequencing. Pedigree analysis of samples collected from her family members revealed that one of her parent and sibling carried the same heterozygous variant but never presented with any disease manifestations most likely due to lack of aPL (data not shown, please contact the corresponding author to request access to data.).

**Figure 2.**
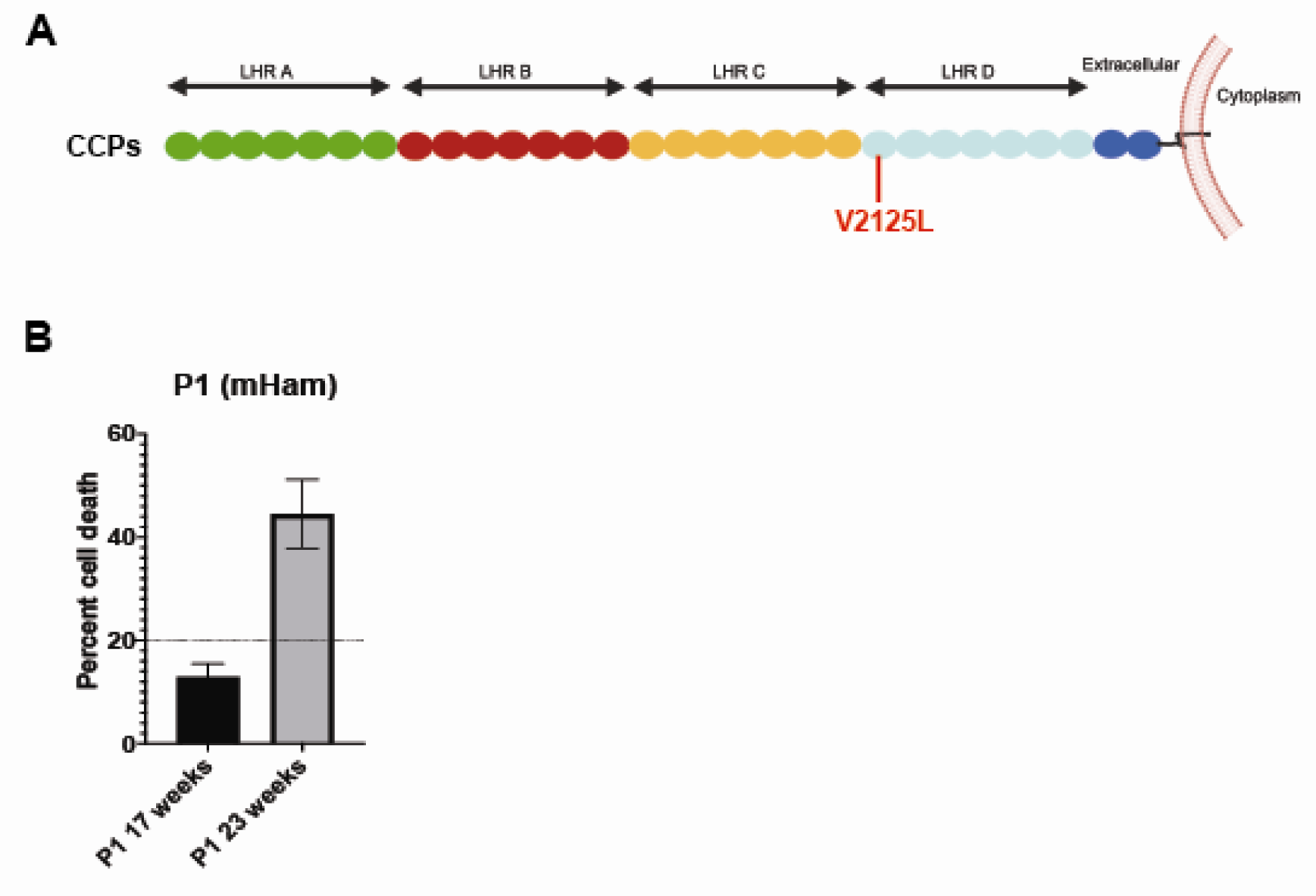
Novel Complement Receptor 1 (CR1) variants and Pedigree tree. A. Overview of CR1 protein, which contains 4 long homologous regions (LHR A, B, C and D). Each LHR consists of 7 complement control proteins (CCPs). *CR1* variant V2125L was identified in CCP22 **B.** modified Ham (mHam) assay demonstrated complement-mediated cell death in Patient 1 (P1) at 23 weeks compared to the 17-week sample. The dashed line represents the threshold for a positive mHam assay.

Missense variants in *CR1* were identified in 3 additional patients (Table 3 and S2). We generated two knock-in cell line models with the CAPS patient-specific variants (V2125L from P1 and S1982G from PD1; Figure S1). We also identified the *CR1-*G2109S variant (*CR1*^G2109S^; negative control) in a woman without APS who presented with pre-eclampsia (separate cohort). These variants were studied because they were located either within the same exon (G2109S) or in adjacent exons (S1982G, Figure 3A).

**Figure 3.**
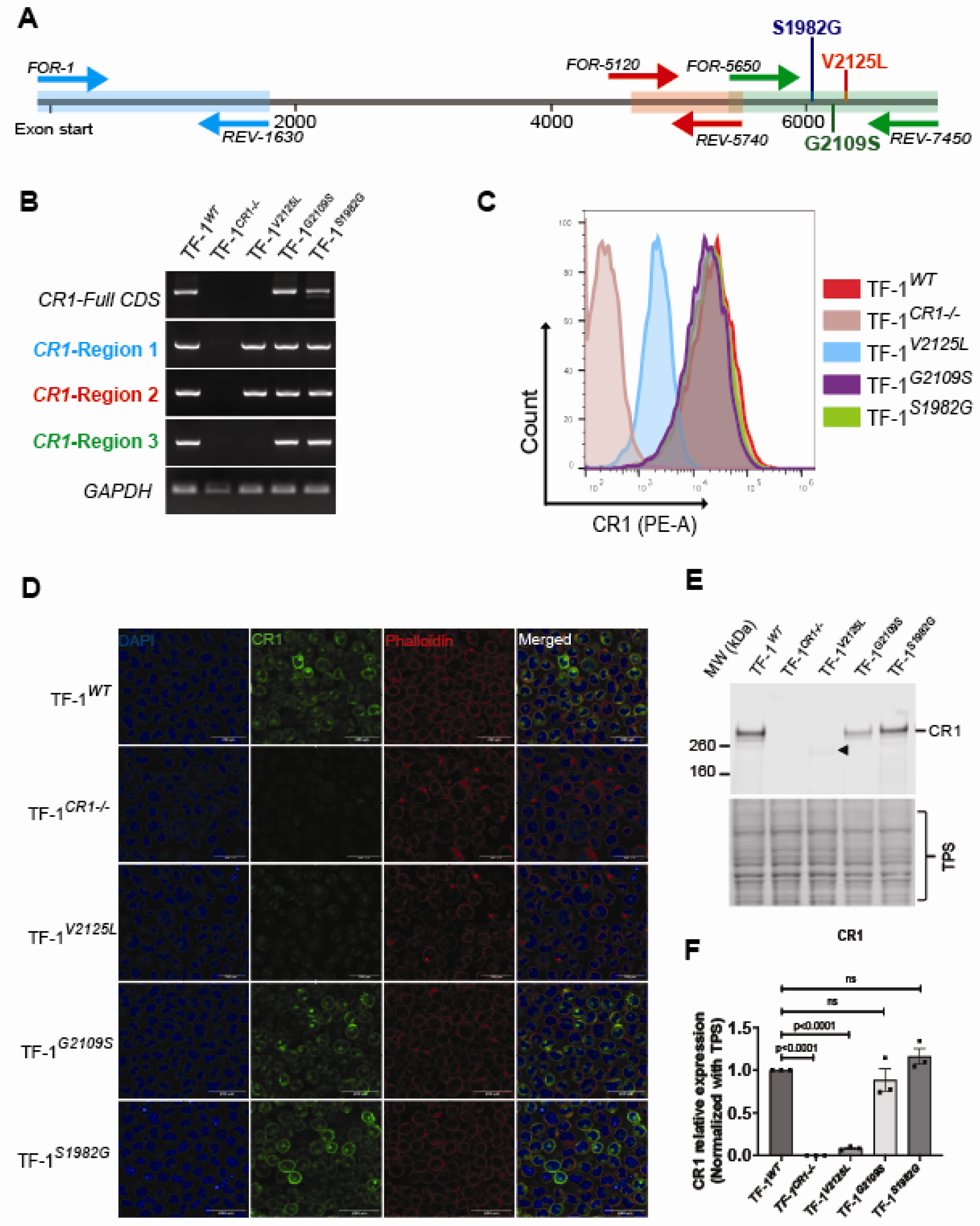
V2125L abolishes CR1 expression. **A.** Representation of *CR1* coding sequence (CDS). CDS was PCR amplified from three regions: region 1(blue), region 2 (red), and region 3 (green) to analyze the role of variants of unknown significance. Specific primer sets are described in Table S2. **B.** Isolated RNA from respective cells was subjected to RT-PCR for cDNA synthesis. cDNA templates were used for semi-quantitative PCR demonstrating that the V2125L variant results in failure to fully transcribe *CR1,* whereas variants G2109S and S1982G did not affect *CR1* transcription. **C.** Flow cytometry analysis represents CR1 expression on respective cell surfaces TF-1*^WT^,* TF-1*^CR1-/-^*, TF-1*^V2125L^*, TF-1*^G2109S^ and* TF-1*^S1982G^*. **D.** Immunofluorescence staining shows that CR1 expression (Green) is significantly reduced in TF-1*^V2125L^*. Nuclei are stained with DAPI (blue) and Phalloidin (red) was used to stain the actin cytoskeleton. The images were captured using a Leica SP8 confocal microscope. The scale bar in the figure insets represents 200µm. **E.** Western blotting represents CR1 protein expression in TF-1*^WT^*, TF-1*^CR1-/-^,* TF-1*^V2125L^,* TF-1*^G2109S^* and TF-1*^S1982G^.* Black arrowhead represents a possible allotypic variant caused by the V2125L mutation. A total protein stain was used to verify equal loading of proteins. **F.** Densitometric quantification of relative CR1 expression through western blot. Images representative of n=3 experiments, data is presented as mean ± SEM; ns = not significant; **** = P < 0.0001.

**Table 2.** Clinical feature of APS patient cohort.

| Patient identification | Age (y)/ Sex | Abs profile | TE history | CR1 % expression |
| --- | --- | --- | --- | --- |
| APS1 | 21-25, M | LAC, secondary | Myocardial infarction | 76.3 |
| APS2 | 66-70, F | LAC, primary | PE, DVT | 74.1 |
| APS3 | 51-55, M | LAC, primary | Arterial thrombosis (necrotic toes), thrombocytopenia | 85.4 |
| APS4 | 31-35, F | aCL IgM<br>aβ2GPI IgM*, primary | Stroke | 85.2 |
| APS5 | 36-40, F | aCL IgM*, primary | Late fetal loss | 88.9 |
| APS6 | 41-45, F | Triple positive, primary | Recurrent DVT and PE | 92.4 |
| APS7 | 36-40, M | aβ2GPI IgG, primary | Bilateral retinal vein occlusion, myocardial infarction | 82.2 |
| APS8 | 66-70, F | LAC, aCL IgM,<br>secondary | PE | 97.9 |

LAC, Lupus anticoagulant; aCL, Anti-cardiolipin antibodies; a β2GPI, Anti β2 Glycoprotein-I antibodies; PE, Pulmonary Embolism; DVT, Deep Vein Thrombosis
\* Meets International Society on Thrombosis and Hemostasis criteria for APS, not ACR/EULAR. All other patients meet both criteria.

**Table 3.** *CR1* Genotyping of CAPS patients.

| Sample | Previously reported polymorphisms |  |  | Novel polymorphisms |  |  |
| --- | --- | --- | --- | --- | --- | --- |
|  | SNV: A3650G (rs2274567) | SNV: HindIII (rs11118133) | SNV: GATA1 (rs11117991) | SNV: V2125L (rs202148801) | SNV: H1503G (rs758055423) | CR1 % expression |
| P1 | Absent | Absent | Absent | Present | Absent | 15.2 |
| P2 | Present | Present | Present | Absent | Absent | 8.4 |
| P3 | Present | Present | Present | Absent | Absent | 1.9 |
| P4 | Present | Present | Absent | Absent | Absent | 23 |
| P5 | Absent | Absent | Absent | Absent | Absent | 20.9 |
| P6 | NS | NS | NS | NS | NS | 27.2 |
| P7 | Present | Present | Absent | Absent | Absent | 15.9 |
| P8 | Absent | Absent | Absent | Absent | Present | 56.3 |
| P9 | Absent | Absent | Absent | Absent | Absent | 27.4 |
| PD1<sup>†</sup> | NS | NS | NS | NS | NS | - |
| PD2<sup>†</sup> | NS | NS | NS | NS | NS | - |

NS Not sequenced
<sup>†</sup> Deceased
\* TF-1 knock-in (KI) cell lines were generated, because these were the first identified CR1 variants.

### *CR1-*V2125L (rs202148801) abolishes *CR1* mRNA and protein expression

We evaluated *CR1* expression in the respective cell lines (TF-1*^WT^*, TF-1*^CR1-/-^*, TF-1*^V2125L^*, TF-1*^G2109S^* and TF-1*^S1982G^*) cells. To evaluate the role of the variant, the coding sequence (CDS) was PCR amplified from three regions using specific primers (Figure 3A, Table S3). We found that the V2125L mutation abolishes *CR1* full-length mRNA expression to levels consistent with the CR1 KO, whereas the G2109S and S1982G variants did not affect *CR1* transcription (Figure 3B). Flow cytometry, immunofluorescence and western blot analysis also confirmed that surface expression of CR1 was reduced in TF-1*^V2125L^* cells when compared to the TF-1*^WT^*, TF-1*^G2109S^* and TF-1*^S1982G^* (Figure 3C-F). Western blot analysis revealed a low molecular weight (black arrow in Figure 3E) CR1 protein in TF-1*^V2125L^* which suggests the V2125L polymorphism yields an allotypic size variant, either by aberrant mRNA splicing or unequal crossing over and loss in the LHR, as described previously [38].

To determine whether the V2125L variant in *CR1* induces protein misfolding and subsequent proteasomal degradation, potentially leading to CR1 loss, we treated TF-1*^WT^* and TF-1*^V2125L^* cells with the proteasome inhibitor bortezomib (BTZ). However, CR1 expression levels remain unchanged in TF-1*^V2125L^* (Figure S4).

### Polymorphisms and hypermethylation resulting in CR1 downregulation

Polymorphisms on *CR1*, rs2274567: G, *Hind*III on intron 27 rs11118133: T and rs11117991:C, have been previously reported to reduce CR1 expression and are associated with the Helgeson phenotype. We performed targeted sequencing and investigated these previously reported polymorphisms in our CAPS patient cohort and found that P2 and P3 had all three Helgeson variants whereas P1, who possessed the novel germline variant, *CR1* SNV rs202148801 (V2125L) linked to low CR1 expression, lacked the previously identified Helgeson variants (Table 3). Two patients, despite low erythrocyte CR1, lacked all known *CR1* polymorphisms (Table 3). We next investigated the methylation profile of *CR1* gene in healthy controls and the CAPS cohort. All CAPS patients except (P1) displayed a hypermethylated profile in the CR1 promoter region as compared to controls (Figure 4A, 4B), suggesting transcriptional silencing as an alternative mechanism for CR1 regulation. To investigate further, CR1 expression across hematopoietic cells, lymphocytes and neutrophils were also analyzed since they too express CR1. In erythroid cells, CR1 expression is primarily regulated by GATA1, whereas PU.1 plays a dominant role in development and function of neutrophils and lymphocytes[16].

**Figure 4.**
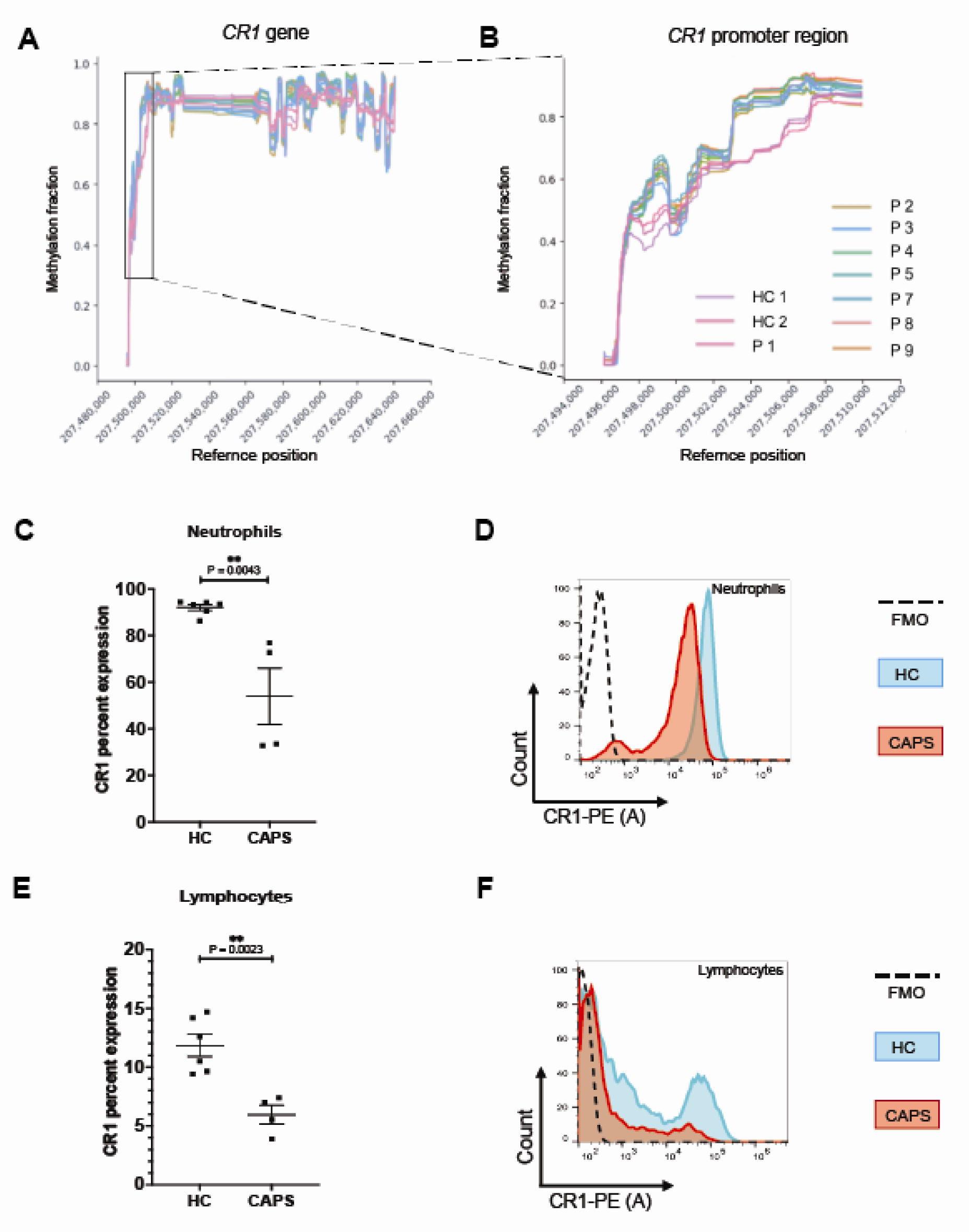
Hypermethylation results in *CR1* downregulation. **A.** Representation of methylation fraction profile of *CR1* gene on the X-axis and with the reference position on the Y-axis. **B.** Inset of the methylation fraction around *CR1* promoter region in healthy controls and CAPS patients. Showing hypermethylation in CAPS patients except index patient (P1) with V2125L SNV as compared to the healthy controls. **C-F**. CR1 percent expression was assessed by flow cytometry in **C.** neutrophils and **E.** lymphocytes of healthy controls (n=6) and CAPS patients (n=4). Live cells were gated using Live/Dead dye, CD45 staining was used to gate the lymphocytes and CD15 staining was used to gate neutrophils. The neutrophils with CR1 on the cell surface were determined by CD15+ CR1+ cell population. Fluorescence Minus One (FMO) controls were used to determine the positive threshold and define the background signal. **D**. Example histogram of CR1 expression in neutrophils, and **F.** Lymphocytes. Data is represented as mean ± SEM and P values are calculated using a two-tailed student t-test, ** = P < 0.01.

We observed significant loss of CR1 in neutrophils (n=4, 53.99 ± 12.05) and lymphocytes (n=4, 5.96 ± 0.79) in the CAPS cohort as compared to healthy control neutrophils (n=6, 92.06 ± 1.28) and lymphocytes (n=6, 11.84 ± 0.94; Figure 4C-4F). Taken together, these results suggest that transcriptional silencing of CR1 due to hypermethylation in the CR1 promotor region can lead to a Helgeson-like phenotype.

### Low CR1 modulates cell surface complement fragment deposition and increases complement-mediated cell death

We next examined functional consequences of the low CR1 on cell surface. First, we analyzed complement activation and deposition on different cell lines using C3c and C3dg monoclonal antibodies. Following C3b deposition, Factor H in the serum and CR1 can serve as the cofactor for the conversion of C3b to iC3b. Final degradation of iC3b into C3c and C3dg is CR1-dependent [21]. C3b deposition and its cleavage fragments were monitored using flow cytometry with C3c and C3dneo monoclonal antibodies (mAbs). Monoclonal anti-C3c detects uncleaved C3b and iC3b while monoclonal anti-C3d ab detects C3b, iC3b and C3dg. No C3b cleavage was observed on TF-1*^CR1-/-^* and TF-1*^V2125L^* when compared to TF-1*^WT^*(31.77%), TF-1*^G2109S^* (49.78%) and TF-1*^S1982G^*(44.25%) after 90 mins (Figure 5A, 5B, Figure S3A, Table S4). C3c deposition was significantly higher in cells lacking CR1 (TF-1*^CR1-/-^*and TF-1*^V2125L^*) (Figure S3B).

**Figure 5.**
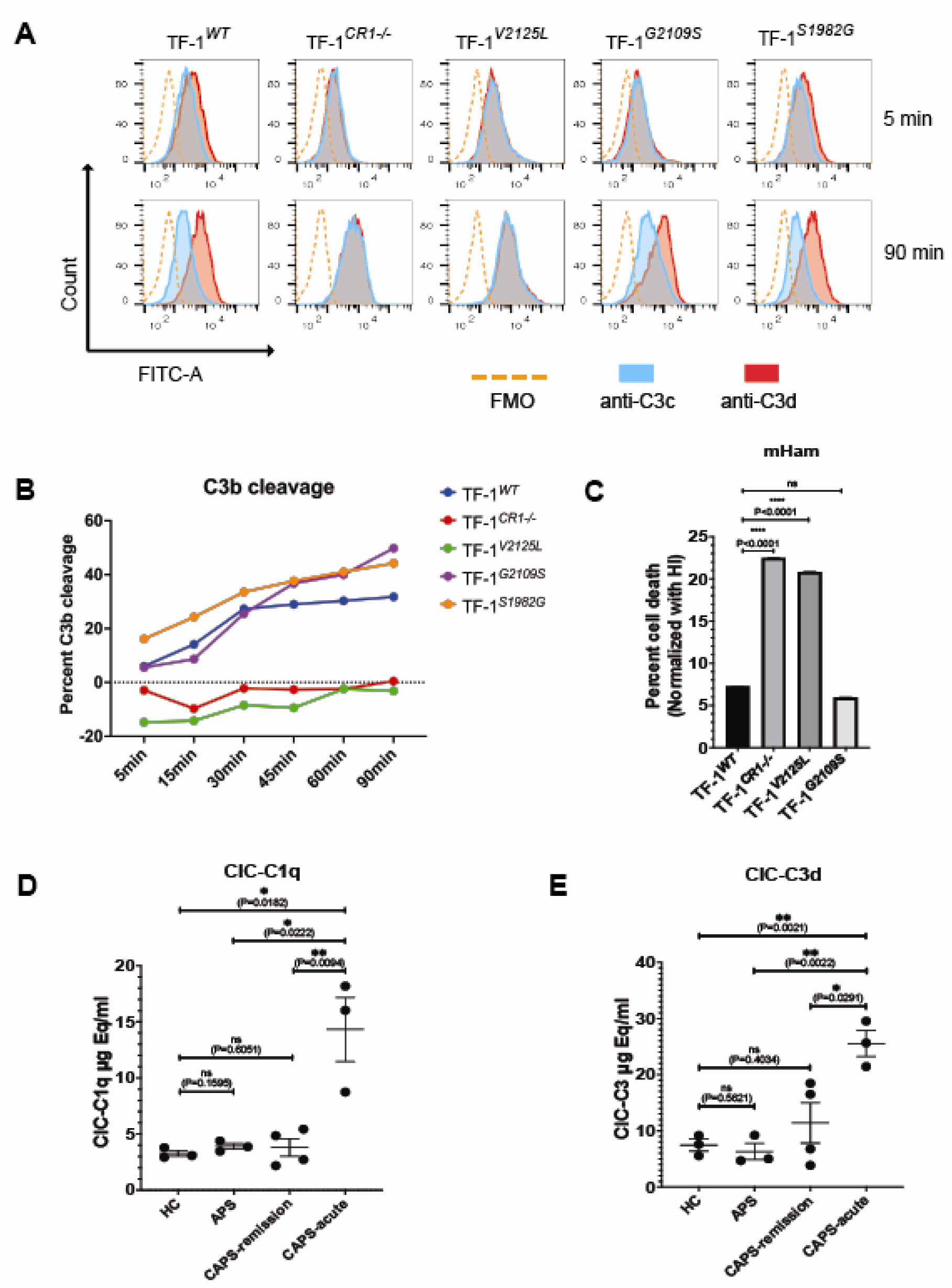
Absence of CR1 leads to complement dysregulation and enhances complement-mediated cell death. **A.** Cell lines were exposed to normal human serum (NHS) for 5-90 mins and surface C3b and its fragments were detected by flow cytometry. Representative images at 5 and 90 mins for each cell line are shown. Monoclonal anti-C3c detects uncleaved C3b and iC3b while monoclonal anti-C3d ab detects C3b, iC3b and C3d, g. FITC labeled goat anti-mouse was used as the secondary ab. Blue solid lines represent C3c deposition and red solid line represents C3d deposition, dashed line represents fluorescence minus one (FMO) control, exposed only to secondary ab. C5 inhibitor was used in the serum to ensure the viability of the cells. **B.** Representative example of C3b cleavage in the cell lines. Time course of C3b cleavage demonstrates the inability of cells to cleave C3b to C3d,g without CR1. Data represents median fluorescence intensity (MFI), representative analysis of n=3 experiments. **C.** modified Ham (mHam) test was performed to analyze complement-mediated cell death in the following cell lines: TF-1*^WT^* (7%), TF-1*^CR1-/-^* (∼23%), TF-1*^V2125L^* (∼21%) and in TF-1*^G2109S^* (∼5%), representative analysis of n = 3 experiments, P value was calculated using two-tailed student t-test. data presented as mean ± SEM; ns= not significant; **** P < 0.001). **D.** Serum specimens (diluted 1:50) from represented cohorts were subjected to ELISA immunoassay to evaluate immune complexes bound to immobilized human C1q purified protein and **E.** ability of immune complexes to covalently bind C3 fragments. The µg Eq/ml concentration for each sample is represented in HC, n=3; APS, n=3; CAPS-remission, n=4; CAPS-acute, n=3. Data is represented as mean ± SEM and P values are calculated using a two-tailed student t-test, ns= not significant; * = P < 0.05; **= P < 0.01.

Further, we performed the mHam assay using the four-cell line to assess the impact of the *CR1*^V2125L^ on complement regulation [12]. TF-1*^V2125L^* cells were as susceptible to complement-mediated cell death induced by NHS as TF-1*^CR1-/-^* cells (∼21% and 23%, respectively), and were more susceptible to complement-mediated killing than TF-1*^WT^* (7%) *and* TF-1*^G2109S^*(5%) cells (Figure 5C).

### Low CR1 increases circulating immune complexes (CIC)

To test the hypothesis that an increase in circulating immune complexes due to low CR1 contributes to increased complement activation, we quantified CIC-C1q ELISA in serum from patients with acute CAPS, CAPS in remission, thrombotic APS and healthy controls. CICs were significantly increased in sera from acute CAPS (CIC-C1q=14.32µg Eq/ml; CIC-C3=25.55 µg Eq/ml) compared to CAPS sera collected in remission (CIC-C1q=3.798 µg Eq/ml and CIC-C3=11.40 µg Eq/ml), healthy controls (CIC-C1q=3.267µg Eq/ml and CIC-C3=7.445 µg Eq/ml), and APS patients (CIC-C1q=3.913 µg Eq/ml and CIC-C3=6.316 µg Eq/ml) (Figure 5D, 5E).

### CAPS is triggered through classical complement activation

We next assessed complement-mediated cell killing induced by CAPS patient sera (remission or acute) using the bio-mHam. Serum samples collected from acute CAPS patients led to higher complement-mediated cell killing of the HEK293*^PIGA-/-^* bioluminescent cells compared to sera from CAPS patients in remission (Figure 6A-B). As compared to their respective serum collected during the acute CAPS episode, subsequent serum samples from five patients treated with therapeutic terminal complement inhibition (C5i), demonstrated significant inhibition of complement-mediated cell death (Figures 6C-D). In vitro addition of the classical pathway inhibitor (sutimlimab) and C5i (eculizumab) but not factor D inhibitor (ACH-5548/ACH-4471) to serum from patients with acute CAPS rescued the cells from complement-mediated cell death, confirming that complement activation in CAPS is predominantly through the classical pathway

**Figure 6.**
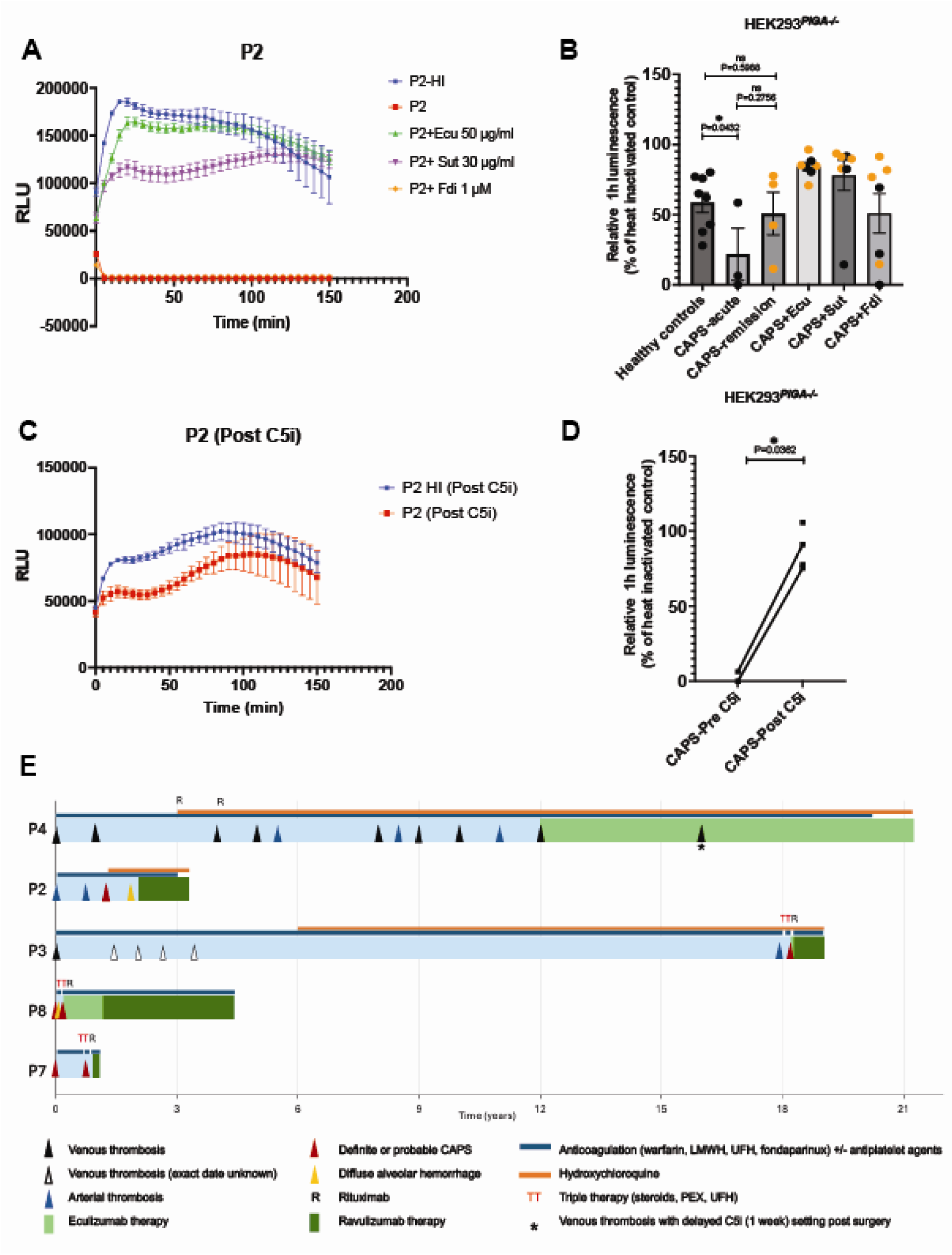
Bioluminescent mHam (bio-mHam) represents enhanced complement activation and efficacy of C5i in CAPS. HEK293*^PIGA-/-^* cells were used for monitoring complement activation in CAPS sera through bio-mHam assay. Relative luminescence units (RLU) were measured every 5 minutes for 2.5 h. **A.** Representative example of CAPS sera on *PIGA*KO cells with or without heat inactivation (HI), eculizumab (Ecu, 50 µg/ml), sutimlimab (Sut, 30 µg/ml) or ACH-5548 (Fdi, 1 µM). **B.** Summary of relative luminescence at 1 h for healthy controls (HC, n= 8), CAPS (acute, n=3 or remission, n=4), with the addition of eculizumab, sutimlimab and Fdi. CAPS remission samples are represented as orange dots. **C.** Representative example to monitor the efficacy of therapeutic C5i in CAPS sera after C5i therapeutic treatment. **D.** Summary of significant reduction of complement-mediated cell killing after C5i therapy in acute CAPS patients. Data is represented as mean ± SEM and P values are calculated using a two-tailed student t-test, ns= not significant: *=p<0.05. **E.** Swimmer plot illustrating the timeline of venous thrombosis, arterial and microvascular thrombotic events and CAPS episodes in patients receiving C5 inhibition (C5i) therapy. Therapy durations for Eculizumab and Ravulizumab are depicted with green and dark green regions, respectively. P2 and P4 are off anticoagulation. LMWH, Low Molecular Weight Heparin; UFH, Unfractionated Heparin; PEX, Plasmapheresis Exchange.

### C5 inhibition mitigates thrombosis in CAPS

Five patients from the CAPS cohort (P2, P3, P4, P7, and P8) were treated with either eculizumab or ravulizumab at the discretion of the treating clinician (duration of therapy 2 months to 9 years). Eculizumab and ravulizumab were administered at the doses recommended for treatment of atypical hemolytic uremic syndrome, at maintenance dosing of 900 mg every 2 weeks for eculizumab and 3300 or 3600 mg ravulizumab every 8 weeks depending on weight. Collectively these patients had over 40 thrombotic events despite therapeutic anticoagulation before therapeutic C5 inhibition. Since starting therapy there has been only one thrombotic event when P4 received her eculizumab 1 week after it was due. Since restarting eculizumab she has had no further thromboses despite discontinuing her anticoagulation over one year ago. The swimmer plot summarizes the thrombotic events and duration of C5i (Figure 6E)

## Discussion

Complement dysregulation in a subset of APS patients predisposes to inflammation and thrombotic events and is a feature of CAPS.[11, 39–41]. Here, we demonstrate that a severe reduction in CR1 expression on erythrocytes, in APS, is a biomarker for severe APS/CAPS. We demonstrate that low CR1 results from novel germline variants in CR1 (CR1V2125L, rs202148801) or hypermethylation of the CR1 promoter region which may occur in the context of previously described CR1 variants associated with the Helgeson phenotype such as rs11117991. Acquired loss of CR1 in the acute phase of CAPS may also result from proteolytic cleavage of immune complexes bound to erythrocyte CR1 by macrophages of the reticuloendothelial system, [33] as is reported in Systemic Lupus Erythematosus (SLE) [17]. However, our finding of reduced CR1 on leukocytes in CAPS suggests that genetic and epigenetic mechanisms play an important role. We show that functionally CR1 deficiency leads to impaired clearance of immune complexes promoting ongoing classical complement activation, as evidenced by an increase in circulating immune complexes and classical pathway activation in sera from patients with acute CAPS. Finally, we demonstrate that complement inhibition at C5 inhibits classical pathway activation *ex vivo* and therapeutic C5 inhibition *in vivo* blocks complement activation and prevents thrombosis in CAPS patients. Together with our previous findings in APS[36], this study supports the potential benefit of complement-targeted therapies to improve patient outcomes and prevent recurrent thrombosis.

We demonstrate a potential association between severe APS/CAPS and the Helgeson-like phenotype (low CR1). Eight out of nine patients in the CAPS cohort met the criteria for the Helgeson-like phenotype on their erythrocytes (Figure 1B), whereas none of the healthy controls or APS patients displayed this phenotype. CR1 expression in CAPS erythrocytes was markedly reduced to an average of 26% of HC levels, closely resembling the Helgeson phenotype (30%), whereas erythrocytes from the APS cohort showed normal expression levels at 103% of the HC mean. The Helgeson phenotype was originally identified serologically, however, serologic evaluation of Knops antigens is often not feasible due to a lack of available ABO-compatible antisera. Additionally, variations in antibody potency and antigen expression between erythrocytes of similar phenotypes may lead to discrepancies when positive reactivity is observed for erythrocytes with low CR1 copy number[28]. Erythrocyte flow cytometry appears to be a simple screening approach to identify APS patients who may be at risk of developing CAPS. We correlated CR1 expression in our CAPS cohort with the already known Helgeson variants and uncovered a novel SNV, *CR1*^V2125L^ (rs202148801) that results in low CR1 expression leading to a Helgeson-like phenotype. Out of eight sequenced CAPS patients (Table 3), six (75%) harbored SNV associated with low CR1 expression and two had rs11117991 (25%), which was demonstrated by Wu et al.,[32] to block GATA1 binding.

GATA1 is primarily recognized for its role in erythroid cells, whereas the transcription factor PU.1 is essential for development and function of myeloid cells (e.g., neutrophils, monocytes) and lymphoid cells[16]. Our findings reveal that hypermethylation in the CR1 promoter region may play a significant role in regulating its expression in CAPS patients. All CAPS patients, except P1, exhibited a similar hypermethylation profile in the CR1 promoter region compared to HC, suggesting epigenetic silencing as a potential mechanism underlying CR1 deficiency. Furthermore, CR1 expression in neutrophils and lymphocytes from CAPS patients was reduced to 58.6% and 50.3% of HC levels, respectively, indicating systemic downregulation across hematopoietic cell types. These observations suggest that CR1 expression is governed not only by genetic predisposition but also by epigenetic modifications, such as promoter hypermethylation, reflecting a dynamic interplay between inherited and environmentally influenced regulatory factors. This hypothesis is further reinforced by the observed increased CR1 expression during clinical remission in CAPS patients with the Helgeson phenotype (P2 and P3, Figure 1C).

Dysregulation in the function of CR1 may amplify classical complement activation by failing to control immune complexes, a known activator of the classical pathway. Indeed, almost 90% of CAPS patients have evidence of classical complement activation by serologic assays compared to roughly 35% of patients with thrombotic APS[36]. Using a patient-specific variant cell line, we characterized the consequences of reduced CR1 expression that lead to increased C3b deposition and enhanced complement-mediated cell death. A major function of CR1 is to bind and process C3b/C4b coated immune complexes[19]. Low CR1 influences immune adherence by reducing the binding of opsonized immune complexes containing C3b and C4b to erythrocytes, leading to an inflammatory response[42–44]. We observed a significantly increased level of immune complexes in the serum of acute CAPS patients as compared to healthy controls and remission CAPS patients.

The etiology of CAPS is multifactorial with genetic and environmental factors contributing to the pathogenesis. Our index case (P1) exemplifies this hypothesis. She inherited the CR1 mutation (*V2125L*) from one of her parent who did not exhibit thrombosis or pregnancy-related complications, most likely because she did not have aPL. Moreover, P1 developed CAPS manifestations only in the setting of pregnancy, a well-established complement amplifying condition, although she had had macrovascular thrombosis before pregnancy. Our study extends the previous case reports and a small series of eculizumab therapy suggesting that C5 inhibition is efficacious in CAPS patients[45, 46]. We extend these observations by identifying two potential biomarkers (erythrocyte CR1 expression and the bio-mHam) to identify patients who are most likely to respond to complement inhibition. Importantly, we show for the first time that it may be feasible to stop anticoagulation in CAPS patients, but this will need validation in larger trials. Two patients (P2 and P4) are off anticoagulation for over 6 and 13 months respectively; thus, similar to paroxysmal nocturnal hemoglobinuria, the prototypic complement-driven thrombophilia, long-term anticoagulation may not be required for patients well-controlled on complement inhibitors[47, 48].

There are limitations in our study. First, the cohort size is small and the follow-up for patients treated with C5i is relatively short. Second, larger cohorts of patients with CAPS and APS are needed to confirm the association between CR1 deficiency and CAPS and the predictive value of the bio-mHam for response to C5 inhibition. Lastly, more patients are needed to confirm the cellular and molecular mechanisms of the Helgeson-like phenotype in CAPS.

In summary, CAPS is a multi-hit disorder of classical complement activation mediated by aPL, impaired immune complex handling, and environmental triggers in the presence of marked CR1 deficiency on erythrocytes. CR1 deficiency exhibits a genetic and epigenetic association with Helgeson-like phenotype, serves as a novel biomarker for CAPS, and is a potential screening tool for risk stratification and response to therapy. The severe suppression of CR1 across multiple cell types (erythrocytes, neutrophils and lymphocytes) in CAPS patients highlights the need for further investigation into therapeutic strategies targeting epigenetic regulation to restore CR1 function. C5i inhibition appears highly efficacious in preventing thrombosis in CAPS, although large multicenter, prospective trials are needed to confirm this finding.

## Data Availability

All data produced in the present study are available upon reasonable request to the authors.

## Acknowledgments

The authors thank Alexion Pharmaceuticals (Boston, MA) for generously providing both Fdi, ACH-4471 and ACH-5548. Graphical illustrations were created with BioRender and Adobe Illustrator. This work was funded by N.H.L.B.I (R56HL133113) and the Department of Defense (W81XWH2110898). The funders had no role in study design, data collection and analysis, the decision to publish or preparation of the manuscript. This work was presented, in part as an oral presentation at the 2024 meeting of the American Society of Hematology in San Diego[49].

## Author Contributions

N.R. designed research, performed research, analyzed data, and wrote the first draft; M.A.C. designed research, performed research, analyzed data, and edited/wrote the paper; G.F.G. designed research and edited/wrote the paper; M.A.C. reviewed research data and wrote/edited the paper; E.M.B. reviewed research data and wrote/edited the paper; D.F.-G. performed research; A.R. performed research; K.H. performed research, reviewed research data and wrote/edited the paper; K.R.M. reviewed data and wrote/edited the paper; S.C. reviewed research data and wrote/edited the paper and R.A.B. designed research, analyzed data, and wrote the paper.

## References

1. Aguiar, C.L. and D. Erkan, Catastrophic antiphospholipid syndrome: how to diagnose a rare but highly fatal disease. Ther Adv Musculoskelet Dis, 2013. 5(6): p. 305–14.

2. Miyakis, S., et al., International consensus statement on an update of the classification criteria for definite antiphospholipid syndrome (APS). J Thromb Haemost, 2006. 4(2): p. 295–306.

3. Cohen, H. and D.A. Isenberg, How I treat anticoagulant-refractory thrombotic antiphospholipid syndrome. Blood. 137(3).

4. Farmer-Boatwright, M.K. and R.A. Roubey, Venous thrombosis in the antiphospholipid syndrome. Arterioscler Thromb Vasc Biol, 2009. 29(3): p. 321–5.

5. Crowther, M.A., et al., A Comparison of Two Intensities of Warfarin for the Prevention of Recurrent Thrombosis in Patients with the Antiphospholipid Antibody Syndrome. N Engl J Med, 2003. 349: p. 1133–8.

6. Nayer, A. and L.M. Ortega, Catastrophic antiphospholipid syndrome: a clinical review. J Nephropathol, 2014. 3(1): p. 9–17.

7. Agostinis, C., et al., A non-complement-fixing antibody to beta2 glycoprotein I as a novel therapy for antiphospholipid syndrome. Blood, 2014. 123(22): p. 3478–87.

8. Carrera-Marin, A., et al., C6 knock-out mice are protected from thrombophilia mediated by antiphospholipid antibodies. Lupus, 2012. 21(14): p. 1497–505.

9. Fischetti, F., et al., Thrombus formation induced by antibodies to beta2-glycoprotein I is complement dependent and requires a priming factor. Blood, 2005. 106(7): p. 2340–6.

10. Pierangeli, S.S., et al., Requirement of activation of complement C3 and C5 for antiphospholipid antibody-mediated thrombophilia. Arthritis Rheum, 2005. 52(7): p. 2120–4.

11. Chaturvedi, S., E.M. Braunstein, and R.A. Brodsky, Antiphospholipid syndrome: Complement activation, complement gene mutations, and therapeutic implications. J Thromb Haemost, 2021. 19(3): p. 607–616.

12. Gavriilaki, E., et al., Modified Ham test for atypical hemolytic uremic syndrome. Blood, 2015. 125(23): p. 3637–46.

13. Maga, T.K., et al., Mutations in alternative pathway complement proteins in American patients with atypical hemolytic uremic syndrome. Hum Mutat, 2010. 31(6): p. E1445–60.

14. Osborne, A.J., et al., Statistical Validation of Rare Complement Variants Provides Insights into the Molecular Basis of Atypical Hemolytic Uremic Syndrome and C3 Glomerulopathy. J Immunol, 2018. 200(7): p. 2464–2478.

15. Fearon, D.T., Identification of the membrane glycoprotein that is the C3b receptor of the human erythrocyte, polymorphonuclear leukocyte, B lymphocyte, and monocyte. J. Exp. Med, 1980. 152 (1).

16. Anderson, K.L., et al., Neutrophils Deficient in PU.1 Do Not Terminally Differentiate or Become Functionally Competent. Blood, 1998. 92(5): p. 1576–1585.

17. Wilson, J.G., et al., Decreased expression of the C3b/C4b receptor (CR1) and the C3d receptor (CR2) on B lymphocytes and of CR1 on neutrophils of patients with systemic lupus erythematosus. Arthritis Rheum, 1986. 29(6): p. 739–47.

18. Eriksson, O., et al., The Human Platelet as an Innate Immune Cell: Interactions Between Activated Platelets and the Complement System. Front Immunol, 2019. 10: p. 1590.

19. Khera, R. and N. Das, Complement Receptor 1: Disease associations and therapeutic implications. Molecular Immunology, 2009. 46(5): p. 761–772.

20. Erdei, A., et al., New aspects in the regulation of human B cell functions by complement receptors CR1, CR2, CR3 and CR4. Immunol Lett, 2021. 237: p. 42–57.

21. Java, A., et al., Role of complement receptor 1 (CR1; CD35) on epithelial cells: A model for understanding complement-mediated damage in the kidney. Mol Immunol, 2015. 67(2 Pt B): p. 584–95.

22. Vandendriessche, S., et al., Complement Receptors and Their Role in Leukocyte Recruitment and Phagocytosis. Front Cell Dev Biol, 2021. 9: p. 624025.

23. Hourcade, D.E., L.M. Mitchell, and M.E. Medof, Decay acceleration of the complement alternative pathway C3 convertase Immunopharmacology, 1999. 42: p. 167–173.

24. Kremlitzka, M., et al., Complement receptor type 1 (CR1, CD35) is a potent inhibitor of B-cell functions in rheumatoid arthritis patients. Int Immunol, 2013. 25(1): p. 25–33.

25. Oliveira, L.C., et al., Complement Receptor 1 (CR1, CD35) Polymorphisms and Soluble CR1: A Proposed Anti-inflammatory Role to Quench the Fire of "Fogo Selvagem" Pemphigus Foliaceus. Front Immunol, 2019. 10: p. 2585.

26. James G. Wilson, E.E.M., Winnie W. Wong, Lloyd B. Klickstein,* John H. WeisS, and Douglas T. Fearon, Identification of a restriction fragment length polymorphismby a CR1 cDNA that correlates with the number of CR1 on erythrocytes. J. Exp. Med., 1986. 186.

27. *International Society of Blood Transfusion Working Group on Red Cell Immunogenetics and Blood Group Terminology: Blood Group Allele Tables*. Amsterdam: ISBT, 2024.

28. Moulds, J.M., et al., Antiglobulin testing for CR1-related (Knops/McCoy/Swain-Langley/York) blood group antigens: negative and weak reactions are caused by variable expression of CR1. Vox Sang, 1992. 62(4): p. 230–5.

29. Herrera, A.H., et al., Analysis of complement receptor type 1 (CR1) expression on erythrocytes and of CR1 allelic markers in Caucasian and African American populations. Clin Immunol Immunopathol, 1998. 87(2): p. 176–83.

30. Zorzetto, M., et al., Complement receptor 1 gene polymorphisms in sarcoidosis. Am J Respir Cell Mol Biol, 2002. 27(1): p. 17–23.

31. Cooling, L., Blood Groups in Infection and Host Susceptibility. Clin Microbiol Rev, 2015. 28(3): p. 801–70.

32. Wu, P.C., et al., Elucidation of the low-expressing erythroid CR1 phenotype by bioinformatic mining of the GATA1-driven blood-group regulome. Nat Commun, 2023. 14(1): p. 5001.

33. Gutierrez, L., et al., Regulation of GATA1 levels in erythropoiesis. IUBMB Life, 2020. 72(1): p. 89–105.

34. Pevny, L., et al., Erythroid differentiation in chimaeric mice blocked by a targeted mutation in the gene for transcription factor GATA-1. Nature, 1991. 349(6306): p. 257–60.

35. Barbhaiya, M., et al., 2023 *ACR/EULAR antiphospholipid syndrome classification criteria*. Ann Rheum Dis, 2023. 82(10): p. 1258–1270.

36. Chaturvedi, S., et al., Complement activity and complement regulatory gene mutations are associated with thrombosis in APS and CAPS. Blood, 2020. 135(4).

37. Pineton de Chambrun, M., et al., CAPS criteria fail to identify most severely-ill thrombotic antiphospholipid syndrome patients requiring intensive care unit admission. J Autoimmun, 2019. 103: p. 102292.

38. Wong, W.W. and S.A. Farrell, Proposed structure of the F’ allotype of human CR1. Loss of a C3b binding site may be associated with altered function. The Journal of Immunology, 1991. 146(2): p. 656–662.

39. Veronica Venturelli, B.M., Ibrahim Tohidi-Esfahani, David A. Isenberg and H.C.a.M. Efthymiou, Can complement activation be the missing link in antiphospholipid syndrome? Rheumatology, 2024. 00: p. 1–12.

40. Martinez-Flores JA, S.M., Perez D, Lora D, Paz-Artal E, Morales JM, and Serrano A., *Detection of circulating immune complexes of human IgA and* β *2 glycoprotein I in patients with antiphospholipid syndrome symptomatology*. J Immunol Methods, 2015. 422:51–8.

41. Martinez-Flores, J.A., et al., Circulating Immune Complexes of IgA Bound to Beta 2 Glycoprotein are Strongly Associated with the Occurrence of Acute Thrombotic Events. J Atheroscler Thromb, 2016. 23(10): p. 1242–1253.

42. Smith, B.O., et al., Structure of the C3b Binding Site of CR1 (CD35), the Immune Adherence Receptor. Cell, 2002. 108: p. 769–780.

43. Noris, M. and G. Remuzzi, Overview of complement activation and regulation. Semin Nephrol, 2013. 33(6): p. 479–92.

44. Kisserli, A., et al., Acquired decrease of the C3b/C4b receptor (CR1, CD35) and increased C4d deposits on erythrocytes from ICU COVID-19 patients. Immunobiology, 2021. 226(3): p. 152093.

45. López-Benjume, B., et al., Eculizumab use in catastrophic antiphospholipid syndrome (CAPS): Descriptive analysis from the “CAPS Registry”. Autoimmunity Reviews, 2022. 21.

46. Faguer, S. and D. Ribes, Early use of eculizumab for catastrophic antiphospholipid syndrome. Br J Haematol, 2022. 196(2): p. e12–e14.

47. Bodo, I., et al., Complement Inhibition in Paroxysmal Nocturnal Hemoglobinuria (PNH): A Systematic Review and Expert Opinion from Central Europe on Special Patient Populations. Adv Ther, 2023. 40(6): p. 2752–2772.

48. Gerber, G.F., et al., A 15-year, single institution experience of anticoagulation management in paroxysmal nocturnal hemoglobinuria patients on terminal complement inhibition with history of thromboembolism. Am J Hematol, 2022. 97(2): p. E59–E62.

49. Nikhil Ranjan, et al., Genetic or Epigenetic CR1 Deficiency Defines Catastrophic Antiphospholipid Syndrome (CAPS) and Response to Complement Inhibition. Blood, 2024. 144(65).

