## Supplementary data for "Genetic and Epigenetic Dysregulation of CR1 is Associated with Catastrophic Antiphospholipid Syndrome (CAPS)"

**Supplemental data**

**CR1 Deficiency Contributes to Catastrophic Antiphospholipid Syndrome (CAPS): Implications for Diagnosis and Treatment**

**Nikhil Ranjan, PhD**^1^, Michael Cole, MD, DPhil^1^, Gloria F. Gerber, MD^1^, Mark A. Crowther, MD, MSc, FRSC, FRCPC, LLM^2^, Evan M. Braunstein, MD, PhD^1^, Daniel Flores-Guerrero^1^, Kathy Haddaway, MLS (ASCP), SBB (ASCP)^3^, Alexis Reed, RN^1^, Michael B. Streiff, MD^1^, MD Keith R. McCrae, MD^4^, Michelle Petri, MD, MPH^5^, Shruti Chaturvedi, MBBS^1^, and Robert A. Brodsky, MD^1, *^

^1^Division of Hematology, Department of Medicine, Johns Hopkins University School of Medicine, Baltimore, MD, USA

^2^Department of Medicine, McMaster University, Hamilton, ON, Canada

^3^The Johns Hopkins Hospital, Transfusion Medicine Division, Baltimore, MD, USA

^4^Departments of Hematology-Oncology, and Cardiovascular and Metabolic Sciences, Lerner Research Institute, Cleveland Clinic, Cleveland, OH, USA

^5^Division of Rheumatology, Department of Medicine, Johns Hopkins University School of Medicine, Baltimore, MD, USA

**Targeted sequencing and CRISPR Cas9-mediated gene editing**

Blood was collected by venipuncture in EDTA and serum separation tubes. Serum was allowed to clot prior to centrifugation at 4°C then separated and stored at −80°C. Whole blood was used to isolate genomic DNA for targeted gene sequencing. Genomic DNA was isolated using a DNeasy Blood & Tissue Kit (Qiagen, Germany) and quantified with Qubit fluorometric assay. 50ng of DNA was used for library preparation using a custom-seq panel of 24 complement regulatory genes (supplementary table S2, Illumina, USA) according to manufacturer’s guidelines. Briefly, targets were amplified using a Veriti 96-well Thermal Cycler, followed by amplicon digestion, index ligation and purification. Amplicons were then amplified and purified, followed by quantification via Qubit fluorometric assay. Library quality was assessed using Agilent 2100 Bioanalyzer. Subsequently, libraries were normalized and pooled before sequencing via Illumina MiSeq using v3 (600-cycle) reagents performed by the Genetic Resources Core Facility (GRCF) at Johns Hopkins School of Medicine. MiSeq optimization and quality control were performed by the GRCF, and mean amplicon coverage for all samples was 640x. Analysis of raw sequencing data (FASTQ) was performed using the DNA Amplicon pipeline (v2.1.1) via the Illumina BaseSpace platform. Alignment to (GRCh38/hg38) human genome reference was performed using the banded Smith-Waterman algorithm in the targeted regions. Variant calls were made using an Illumina-developed germline variant caller and filtered using VariantStudio software (v3.0). Variants not passing Illumina’s variant quality filters were excluded, followed by filtering using the following criteria to identify rare germline single nucleotide variants and indels: 1) depth greater than 50X; 2) non-synonymous coding region or splice variants; 3) variant allele frequency between 40% and 60%; 4) minor allele frequency less than 0.005 in any ethnic population in the genome aggregation database (gnomAD, total 141,456 individuals). Large deletions were determined by complete loss of signal for multiple consecutive amplicons. Homozygous deletion of CFHR1 and CFHR3, reported to occur in approximately 2% of the population, was included in our analysis due to its association with CFH antibody formation and association with aHUS. 47 Amplicons were designed to cover all exons of the mentioned genes in Table S2.

To understand the methylation traces on CR1, we performed whole genome sequencing and multiomics using duet multiomics evoC kit (Biomodal, Cambridge, UK). 180 ng of DNA was sonicated in Covaris E220 sonicator set to a target size of 250 bp. Library preparation was performed using manufacturer’s protocol. Sequencing was performed on the Illumina NovaSeq X. Libraries were quantified using the Tapestation D5000 (Agilent) and subsequently diluted to 150 pM for loading on the sequencer. To balance out low cytosine content in deaminated libraries, 10% PhiX was added according to the manufacture’s protocol. All libraries were run in a paired-end set up using 300 cycles in a 151/8/8/151 base-reads setup. A minimum of 1 billion read pairs per human genome sample was obtained to achieve approximately 30X mean coverage. Data was analyzed using Biomodal’s analysis pipeline.

The CRISPR/Cas9 system was used to generate CR1 knock-out (KO) and knock-in (KI) cell lines through homology-directed repair (HDR). Single guide RNA (sgRNA) was used to make knock out lines with the help of Synthego gRNA design tool. The gRNA templates and donor templates were cloned in a HDR donor vector using the DNA builder Gibson assembly (NEB, MA, USA). TF-1 cells were transfected using 4D nucleofection technology followed by single-cell sorting to obtain targeted clones. CR1 expression was assessed by quantitative Real Time-PCR (qRT-PCR), western blot and flow cytometry to compare the expression levels with wild-type cells. Single-cell knock-out and knock-in clones will be screened using Sanger sequencing and protein expression using flow cytometry. The experiments described were performed on five cell lines – TF-1*^WT^*, TF-1*^CR1-/-^*, TF-1*^V2125L^*, TF-1*^G2109S^* and TF-1*^S1982G^* unless mentioned otherwise.

**Reverse transcription and semi-quantitative PCR**

RNA was isolated from TF-1*^WT^*, TF-1*^CR1-/-^*, TF-1*^V2125L^*, TF-1*^G2109S^* and TF-1*^S1982G^* cells, and 250ng of RNA was used for cDNA synthesis using RT Supermix (New England Biolabs, MA, USA). cDNA was further used for semiquantitative PCR using KOD one PCR master mix (Sigma-Aldrich, MO, USA). Table S2 describes primers used for coding sequence amplification, cycling conditions for the reactions were as follows: 95^o^C for 30 sec, 35 cycles at 95^o^C for 10 sec, 60^o^C for 5 sec/kb and 68^o^C for 10 sec. Relative expression was quantified using GAPDH as a loading control.

**Western blot**

Whole-cell lysates were prepared using Radioimmunoprecipitation assay (RIPA) buffer (Sigma-Aldrich, MO, USA) supplemented with HALT protease and phosphatase inhibitors (Thermo, MA, USA) as described previously [1]. Protein was quantitated using a Bicinchoninic Acid (BCA) assay and 10ug of protein was subjected to western blotting in 3-8% polyacrylamide gel. Separated proteins were transferred on the (Polyvinylidene Fluoride) PVDF membrane followed by blocking in 5% milk for 1 hour at room temperature, the membrane was probed with CR1 antibody (1:1,000, Abclonal, MA, USA) overnight at 4^o^C. Further, the membrane is washed four times with TBST and probed with an anti-rabbit secondary antibody (1:10,000, LI-COR, NE, USA) for 1 hour at room temperature. The blot was developed after washing four times with TBST using an Odyssey CLx scanner (LI-COR, NE, USA). Total protein staining was used to ensure equal loading of protein and normalization in all samples.

**Immunofluorescence**

10^6^ cells from each cell line were harvested and washed twice with 1X PBS. Cells were fixed with 4% paraformaldehyde for 15 mins, followed by two 1X PBS washes, the cells were permeabilized using (PBST PBS+0.1% triton X-100) for 15 min followed by two 1X PBS washes. Further, the cells were probed with CR1 mAb (10µg/ml, Invitrogen, MA, USA) overnight at 4oC. Post-incubation cells were washed twice with 1X PBS and incubated in goat anti-rabbit AF-488 secondary ab (4µg/ml, Invitrogen, MA, USA). Imaging was performed using a Leica SP8 confocal microscope using a 20X objective lens, 4 fields from each sample were captured and one representative image was shown. The scale bar in the insets of the respective image represents 200µm.

**CR1 expression by flow cytometry**

50µl of whole blood was diluted in 1450ul of Hanks Balanced Salt Solution (HBSS)+1%BSA (HBSSA) and washed at 500g for 5 min. Washed cells were further suspended in 500ul of HBSSA and 10ul of the homogenous suspension was used for respective staining. The final staining solution (50µl) includes 5ul PE-conjugated CR1 antibody (BD Biosciences, NJ, USA) and HBSSA. Cells were stained for 30 minutes at room temperature, and cells without antibodies were used as a control. Cells were washed twice, resuspended in HBSSA and analyzed by flow cytometry using Cytoflex S (Beckman Coulter, CA, USA).

To measure CR1 surface expression on neutrophils and lymphocytes, erythrocytes were lysed using ACK lysing buffer (Quality Biological, MD, USA) using manufacturers protocol. Cells were further stained with Live/Dead dye, CD45 (Invitrogen, MA, USA), CD15, (BioLegend, CA, USA) and CR1. All samples were processed within 24 hours of collection. Fluorescence minus one (FMO) controls were used to set positivity threshold for the population of interest.

**Helgeson phenotype screening**

Red Blood Cells (RBC) samples of Helgeson phenotype (Kn(a-) McC(a-)) were obtained from an internal collection of frozen patient-sourced RBC prepared as previously described [2]. Antisera known to contain anti-Kn^a^, anti-McC^a^ and anti-Yk^a^ were obtained from an internal collection of frozen patient-sourced plasma stored at ≤ -65°C. Due to limited inventory, only one plasma example of each antibody was tested. Additional antibody specificities were not available. Knops antigen-positive RBC (Kn(a+) McC(a+) Yk(a+)) were sourced from commercially available reagent RBC (Immucor, Norcross, GA).

Frozen RBC samples were thawed in RBC storage solution containing 2 drops of 22% albumin warmed to 37°C (Immucor, Norcross, GA). After thawing, the RBCs were manually washed 3 times in saline. RBCs from CAPS patients were manually washed 3 times in saline, and a direct antiglobulin test (DAT) was performed by standard tube method using commercially available anti-IgG (Immucor, Norcross, GA). All RBC examples to be tested, including controls, were resuspended to a 0.8% suspension in diluent (Ortho Clinical Diagnostics, Raritan, NJ). Serologic expression of Kn^a^, McC^a^, and Yk^a^ was determined using the column agglutination method per the manufacturer’s instructions (Ortho Clinical Diagnostics, Raritan, NJ). In summary, 50µL of 0.8% RBC suspension was incubated with 25µL of antisera at 37°C for 15 minutes followed by a 10-minute centrifugation in a calibrated centrifuge (Ortho Clinical Diagnostics, Raritan, NJ).

Results were read macroscopically for presence of agglutination in the gel column. Absence of agglutination indicated absence of the antigen. The presence of agglutination was graded from 1+ to 4+ based on strength of reactivity and indicated a positive result for the antigen being tested. Knops antigen positive (positive control) and Helgeson phenotype RBC (negative controls) were tested with each antiserum and valid results were determined by expected reactions of antisera with positive and negative controls.

**Modified Ham (mHam)**

mHam assay was performed with TF-1*^WT^*, TF-1*^CR1-/-^*, TF-1*^V2125L^*, and TF-1*^G2109S^* cells, as described previously [3]. 7000 cells/well were washed with PBS and resuspended in gelatin veronal buffer (GVB++, 80 µl/well). Pooled normal human serum (NHS) was treated as indicated with heat inactivation (HI) for 30 minutes at 56^o^C to inactivate complement proteins or addition of targeted complement inhibitors. Cells were incubated with 20µl NHS for 45 mins at 37^o^C then washed with PBS. Cells incubated with HI serum were used as control. Next, cells were incubated with WST-1 viability dye for 2 hours and absorbance 450nm and 630nm (A_450nm_-A_630nm_) was measured.

**Bioluminescent mHam (bio-mHam)**

Bio-mHam was performed as described previously[4]. Briefly, bioluminescent HEK293*^PIGA-/-^* cells (490 BioTech) were harvested with Trypsin-EDTA, washed twice with 1X PBS, 40,000 cells/well resuspended in 80µl/well GVB++ buffer. Serum (20µl) was added to initiate the assay after respective treatment (heat inactivation and complement inhibitors). Luminescence was monitored by serial measurements (every 5 min) using a BMG Clariostar luminometer (Ortenberg, Germany) and percent relative luminescence at 1 hour was calculated as the luminescence of cells treated with the patient serum compared to cells treated with the sample’s heat-inactivated control. A relative 1 h luminescence of ~12% was used to establish a “positive” threshold for abnormal complement activity, corresponding to the lower 85^th^ percentile.

**Evaluation of circulating immune complexes (CIC)**

To quantify circulating immune complexes (CIC), CIC-C1q and CIC-C3d (Quidel, San Diego, CA) ELISA immunoassay (EIA) assay was performed. CIC-C1q EIA quantifies the CIC bound to the immobilized human C1q purified protein. Whereas, CIC-C3d EIA quantifies the ability of CR1 receptors to clear the covalently bound immune complexes to the third complement protein (C3). The CIC- containing C3 fragments are captured by using a mAb that specifically binds the iC3b, C3dg and C3d activation fragments of C3. The manufacturer’s protocol was followed. Briefly, participant serum was diluted to 1: 50 using the specimen diluent and 100ul of the specimen was subjected to C1q or C3d coated wells in duplicates for 1 h at room temperature. HRP-conjugated mouse anti-human IgG was added to each test well and incubated for 30 min. Finally, an enzyme substrate was added to each test well for 30 mins, after incubation, a reagent was added to stop color development. The optical densities (OD) were measured at 405nm using a BMG Clariostar reader (Ortenberg, Germany). A standard curve is generated using A405-blank values (on the Y-axis) for each standard and the assigned concentration for each standard (along the X-axis). The concentration for each sample is calculated from the standard curve using linear regression analysis (y=mx+C).

**Quantification and statistical analysis**

All experiments were performed thrice unless mentioned otherwise. Statistical comparisons were made using the student’s two-tailed unpaired t-tests using Prism10 (GraphPad, MA, USA). Results are shown as mean ± standard error mean (SEM) and P values of <0.05 are considered significant. Flow cytometry data was analyzed by FlowJo and GraphPad Prism.

**Supplemental table S1. Gene list of targeted sequencing**

|  | **Gene** | **Symbol** |
| --- | --- | --- |
| 1. | Complement factor B | CFB |
| 2. | Complement factor D | CFD |
| 3. | Complement factor H | CFH |
| 4. | Complement factor I | CFI |
| 5. | Complement factor P | CFP |
| 6. | Phosphatidylinositol Glycan Anchor Biosynthesis class A | PIGA |
| 7. | CD55 | CD55 |
| 8. | CD59 | CD59 |
| 9. | CD46 | CD46 |
| 10. | Complement Factor H Related 1 | CFHR1 |
| 11. | Complement Factor H Related 2 | CFHR2 |
| 12. | Complement Factor H Related 3 | CFHR3 |
| 13. | Complement Factor H Related 4 | CFHR4 |
| 14. | Complement Factor H Related 5 | CFHR5 |
| 15. | N-acetylneuraminate Synthase | NANS |
| 16. | N-Acetylneuraminic Acid Phosphatase | NANP |
| 17. | Diacylglycerol Kinase Epsilon | DGKE |
| 18. | Glucosamine (UDP-N-Alcetyl)-2-Epimerase/N-Acetylmannosamine Kinase | GNE |
| 19. | Thrombomodulin | THBD |
| 20. | Complement C3 | C3 |
| 21. | Complement C5 | C5 |
| 22. | Complement receptor 1 | CR1 |
| 23. | ADAM Metallopeptidase with Thrombospondin Type 1 Motif 13 | ADAMSTS13 |
| 24. | Plasminogen | PLG |

**Table S2. *CR1* variants identified in CAPS/ probable CAPS patients.**

| **Patient identification** | **Nucleotide change** |
| --- | --- |
| P1* | 1:207785099/ G > T |
| P2 | 1:207580276/ A > G; 207589899/ A > T; 1:207521012/T > C |
| P3 | 1:207580276/ A > G; 207589899/ A > T; 1:207521012/T > C |
| P4 | 1:207580276/ A > G; 207589899/ A > T |
| P5 | None |
| P6 | NS |
| P7 | 1:207580276/ A > G; 207589899/ A > T |
| P8 | 1-207748997/ T>G |
| P9 | None |
| PD1^†^ * | 1:207782682/ A > G |
| PD2^†^ | 1:207757992/ A > G |

NS Not sequenced

† Deceased

***** TF-1 knock-in (KI) cell lines were generated, because these were the first identified CR1 variants.

**Supplemental table S3. Primers used for semi-quantitative PCR**

| **Gene/Fragment** | **Primer** |
| --- | --- |
| CR1 CDS-FOR1 (Region 1) | AATGCAATGCCCCAGAATGGC |
| CR1 CDS-REV1630 (Region 1) | GACTTGACCAGACCAGGTTATC |
| CR1 CDS-FOR5120 (Region 2) | TGGAGCCCTGAAGCCCCGAG |
| CR1 CDS-REV5750(Region 2) | CCACATGATTTTCGTCTACAGTTGTC |
| CR1 CDS-FOR5650 (Region 3) | GGGAAAATGTTCTCTATCTCCTG |
| CR1 CDS-REV7450 (Region 3) | TTTGTTTGCAGAGTTCGGGG |

**Supplementary table S4. C3b cleavage analysis.**

| **Time (min)** | **TF-1*^WT^*** | **TF-1*^CR1-/-^*** | **TF-1*^V2125L^*** | **TF-1*^G2109S^*** | **TF-1*^S1982G^*** |
| --- | --- | --- | --- | --- | --- |
| 5 | 5.99 | -2.91 | -14.78 | 5.54 | 16.12 |
| 15 | 14.10 | -9.69 | -14.22 | 8.638 | 24.25 |
| 30 | 27.3 | -2.16 | -8.40 | 25.5 | 33.54 |
| 45 | 28.96 | -2.64 | -9.43 | 36.77 | 37.66 |
| 60 | 30.30 | -2.41 | -2.4 | 39.99 | 40.98 |
| 90 | 31.77 | 0.477 | -3.13 | 49.78 | 44.25 |

C3b cleavage was significantly less in the TF-1*^CR1-/-^* and TF-1*^V2125L^* cells as compared to TF-1*^WT^*, TF-1*^G2109S^* or TF-1*^S1982G^*. Anti-C3c mAb detects the uncleaved C3b and iC3b deposited on the cell surface while the anti-C3d Ab identifies the C3dg fragment and its presence in iC3b and C3b. Hence, the ratio of (C3b+iC3b)/(C3b+iC3b+C3dg) is used to determine the amount of C3b cleaved to C3dg in respective cells. Values represent C3b% cleavage from one experiment.

**Supplemental figures**

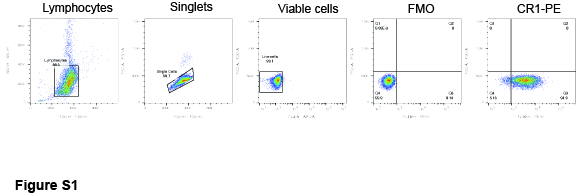

**Figure S1. Gating strategy for CR1 flow cytometry analysis on RBCs.** 50µl of blood from the EDTA tube was resuspended in 1.5ml HBSS + 1% BSA (HBSSA) and centrifuged at 1200rpm for 5min. Supernatant was discarded and pelleted cells were resuspended in 500µl of HBSSA. 10µl of the resuspended samples were used for CR1 staining in a final staining solution of 50µl with 5µl of CR1-PE conjugated antibody. The cells were also incubated with far red live/dead dye, to use the viable cells for the analysis. The RBCs were incubated for 30 mins at room temperature and washed twice with HBSAA. RBCs were resuspended in 300ul HBSAA and used for flow cytometry analysis. Doublets were excluded from the analysis and the viable cells were gated for CR1 expression. A fluorescence minus one (FMO) sample was used as a negative control.

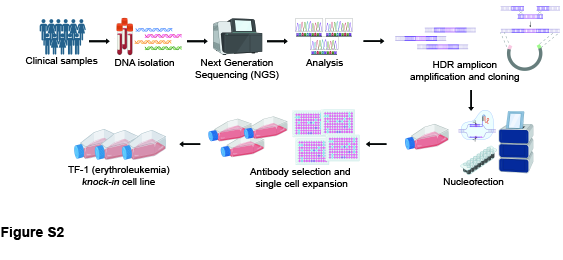

**Figure S1. General procedure for patient specific knock-in cell line generation.** Whole blood samples were obtained in EDTA tubes and 200µl was subjected to DNA isolation. Samples were further used for library preparation using a custom-seq amplicon panel for complement regulatory genes and sent for Illumina MiSeq sequencing. 2nM of each library was pooled together and the sample was used for MiSeq after confirming the quality of the sample using BioAnalyzer, the read length for MiSeq was 2×300. The variants in respective samples were identified. gRNA and HDR amplicons were made according to the locations of respective variants in the genome. TF-1 cells are nucleofected using the gRNA and HDR templates respective patient specific *CR1* variants. Nucleofected cells were transferred to a positive selection media containing puromycin selectable marker and media was changed every 3-4 days. Positively selected cells were single cell sorted in 96 well plate and positive clones with successful knock-in mutation was screened through PCR and Sanger sequencing.

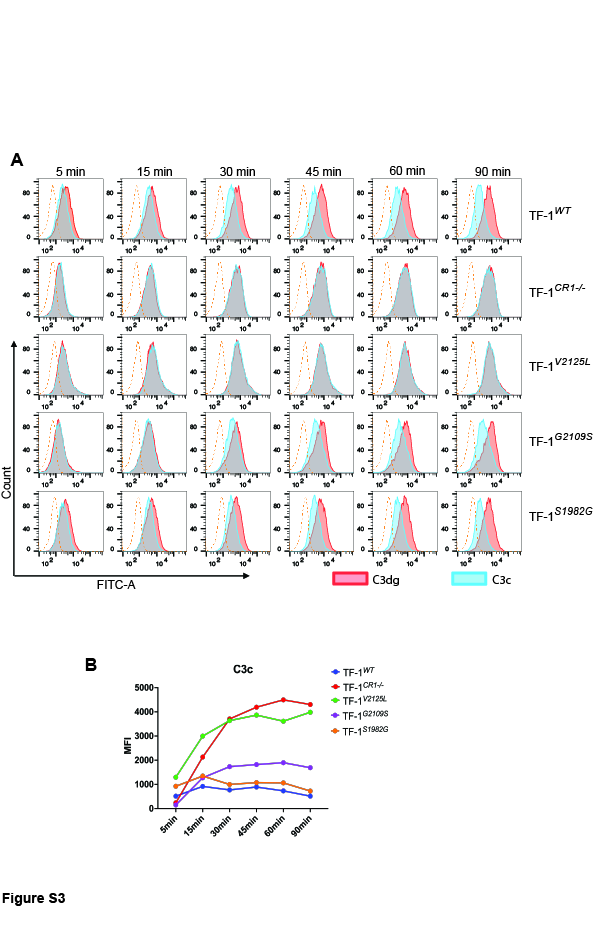

**Figure S3. Absence of CR1 dysregulated complement.** Represented cell lines were exposed to normal human serum (NHS) for 5-90 mins and surface C3b and its fragments were detected by flow cytometry. Representative images at 5 and 90 mins for each cell line is shown. Monoclonal anti-C3c detects uncleaved C3b and iC3b while monoclonal anti-C3d ab detects C3b, iC3b and C3d, g. FITC labelled goat anti-mouse was used as the secondary Ab. Blue solid lines represent C3c deposition and red solid line represents C3d deposition, dashed line represents fluorescence minus one (FMO) control, exposed only to secondary ab. C5 inhibitor was used in the serum to ensure viability of the cells. **B.** Representative example of C3c deposition in various cell lines following incubation with NHS for 5-90 mins.

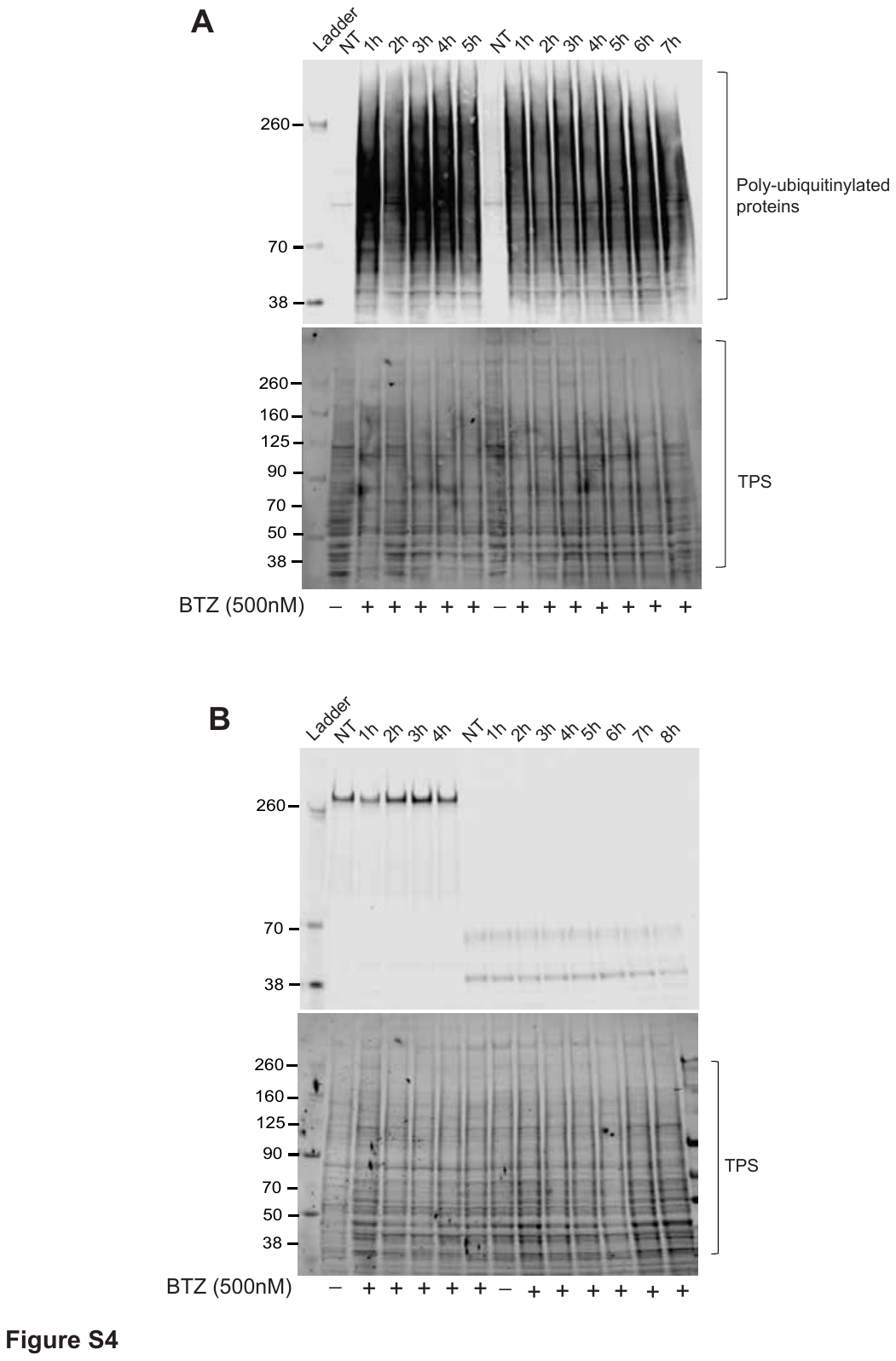

**Figure S4. V2125L mutation does not lead to proteasomal degradation of CR1.** A. TF-1*^WT^* and TF-1*^V2125L^* cells were treated with bortezomib (BTZ; 500nM) for different 1h and samples were collected at mentioned time points to check CR1 expression in respective cell lines through western blotting. The samples were probed with an antibody which recognizes poly-ubiquitinylated proteins, that serves as a positive control for BTZ treatment. Increased amount of poly-ubiquitinylated proteins after BTZ treatment represents proteasome blocking. B. TF-1*^WT^* represents a small increase in CR1 expression after BTZ treatment, whereas the TF-1*^V2125L^* cells do not represent any CR1 expression. Total protein stain (TPS) was used as a loading control for the experiment.

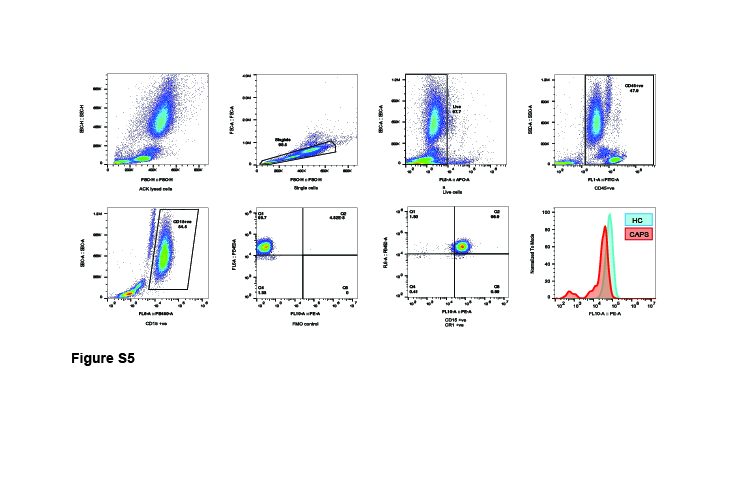

**Figure S5. Gating strategy for CR1 flow cytometry analysis on neutrophils.** Whole blood sample was ACK lysed, washed and resuspended in 1X PBS. Following staining and gating strategy was used: Live/Dead dye was used for viability gating, CD45 stain was used to gate the lymphocytes. For neutrophils identification, highly positive CD15 cells were gated to exclude other granulocytes. A fluorescence minus one (FMO) sample (stain cells with all the markers except protein of interest) was used as a negative control, to set positivity threshold and to define background signal.

**References**

1.	Ranjan, N., et al., *The Tumor Suppressor MTUS1/ATIP1 Modulates Tumor Promotion in Glioma: Association with Epigenetics and DNA Repair.* Cancers (Basel), 2021. **13**(6).

2.	Judd WJ, J.S., and Storry JR, *Judd’s Methods in Immunohematology. 4th ed.* AABB Press, 2022: p. 294-297.

3.	Gavriilaki, E., et al., *Modified Ham test for atypical hemolytic uremic syndrome.* Blood, 2015. **125**(23): p. 3637-46.

4.	Cole, M.A., et al., *Complement Biosensors Identify a Classical Pathway Stimulus in Complement-Mediated Thrombotic Microangiopathy.* Blood, 2024.
